# Dynamics of neurotransmitter and extracellular vesicle-derived microRNA landscapes during heroin and methamphetamine withdrawal

**DOI:** 10.1101/2021.04.19.21255653

**Authors:** Juehua Yu, Fengrong Chen, Yu Xu, Kai Shi, Zunyue Zhang, Qingyan Peng, Zhenrong Xie, Jing Lu, Hongjin Wu, Yuru Ma, Lei Zou, Yong Zhou, Cheng Chen, Jiqing Yang, Yiqun Kuang, Yuan Wang, Tao Tan, Mei Zhu, Trevor W. Robbins, Kunhua Wang

**Affiliations:** NHC Key Laboratory of Drug Addiction Medicine (Kunming Medical University), First Affiliated Hospital of Kunming Medical University, Kunming, Yunnan 650032, China; Centre for Experimental Studies and Research, First Affiliated Hospital of Kunming Medical University, Kunming, Yunnan 650032, China; Yunnan Institute of Digestive Disease, First Affiliated Hospital of Kunming Medical University, Kunming, Yunnan 650032, China; College of Science, Guilin University of Technology, Guilin 541004, China; Department of Psychiatry, First Affiliated Hospital of Kunming Medical University, Kunming, Yunnan 650032, China; Department of R&D, Echo Biotech Co., Ltd, Beijing, China; Institute of Primate Translational Medicine, Kunming University of Science and Technology, Kunming, Yunnan 650500, China; Department of Psychology and the Behavioural and Clinical Neuroscience Institute, University of Cambridge, Cambridge, United Kingdom; Institute of Science and Technology for Brain-Inspired Intelligence, Fudan University, Shanghai, China

**Keywords:** substance use disorders, withdrawal, neurotransmitter, exosomes, microRNA, induced pluripotent stem cells, neurite outgrowth

## Abstract

Circulating miRNAs in small vesicles known as exosomes within blood have been emerging as a new research hotspot in the field of psychiatric disorders. The aim of this work was to characterize the changes in exosomal microRNA profiles, both short-term and long-term, during substance withdrawal using a cross-sectional study design. Using weighted gene co-expression network analysis, a series of known, conserved, and novel exosomal microRNAs were identified as being associated with withdrawal stage and key neurotransmitters GABA, choline, and serotonin. Bioinformatics analyses established that the differences in the miRNA profile target signaling pathways are associated with developmental and intellectual abnormalities. Notably, a set of dysregulated microRNA signatures including hsa-mia-451a and hsa-mir-21a resulted in an AUC of 0.966 and 0.861, respectively, for predicting patients with substance use disorders. Furthermore, hsa-miR-744a-5p was positively correlated with serotonin, and its important role in maintaining neuronal development and function was revealed using an *in vitro* human induced pluripotent stem cells derived neuronal model. Taken together, these data suggest that the microRNA content of circulating exosomes represent a biomolecular “fingerprint” of the progression of substance withdrawal and may uncover the putative mechanism of how these exosomal microRNAs contribute to central nervous system development and function.

## Introduction

According to various estimates, between 35 and 50 million people worldwide suffer from severe substance use disorders (SUDs). These are associated with serious health problems and harmful consequences, including mortality, morbidity, and criminality (Brady, 2019; Lehmann & Fingerhood, 2018). The use of heroin and methamphetamine, the two most commonly abused illegal substances in China, account for approximately 37% and 56.1% of registered drug abusers, respectively (The State Council China, 2019). Both heroin and methamphetamine dependence are chronic, often relapsing brain disorders that cause acute and protracted withdrawal symptoms when their use is discontinued or reduced. For some individuals, severe psychological withdrawal symptoms such as depression, anxiety, reduced motivation, difficulties experiencing pleasure, and apathy can last for weeks, months, or even years. These symptoms may lead patients to seek relief by returning to substance use, resulting in a pattern of repeated relapse (Bell & Strang, 2020; Cruickshank & Dyer, 2009).

In this complex group of disorders which have a multitude of signs and symptoms associated with both nervous system and other body systems, the dysfunctioning of neurotransmitter systems has been proposed to play a significant role in the development of substance dependence (Duman *et al*, 2019; Muller & Homberg, 2015; Tomkins & Sellers, 2001). Increased platelet serotonin was first reported in a small sample of opiate addicts compared with alcoholics and control participants (Schmidt *et al*, 1997). Conversely, a significant increase in plasma dopamine but not platelet serotonin has been shown in heroin and cocaine addicts (Macedo *et al*, 1998). Stimulant abuse (e.g., methamphetamine) is associated with elevated immune activation and lower tryptophan (Carrico *et al*, 2008), in line with the finding that decreased serum tryptophan, a precursor of serotonin, has been found in heroin treated rats (Zheng *et al*, 2013).

Cumulating structural and functional brain changes, as well as neurocognitive deficits and emotional impairments in both heroin and methamphetamine dependent populations have been demonstrated in neuropsychological studies. For example, a general loss of neurons and astrocytes (Buttner & Weis, 2006), a decreased number of neural precursor cells, accompanied by lower rates of proliferation (Bayer *et al*, 2015), and a marked loss of dendritic trees in targeting cells (Oehmichen *et al*, 1996) have been found in the *post mortem* brains of heroin and methamphetamine addicts. These findings suggest that prolonged substance use significantly impacts neurogenesis and the way the brain works by changing the way nerve cells communicate with one another. In animal models, chronic administration of illegal abused substances results in alterations to synaptic plasticity and neuronal apoptosis in the nucleus accumbens, ventral tegmental area, and hippocampus (Kourrich *et al*, 2015; Pignatelli & Bonci, 2015). However, whether the degenerative processes with adverse effects on neurodevelopmental and neuropsychiatric functions associated with substance dependence can be discontinued or reversed through the elimination of addictive substances remains questionable (Norman *et al*, 2009). Only limited molecular biomarkers have been associated with withdrawal symptoms and the mechanisms underlying altered communications between the peripheral circulation and the central nervous system (CNS) that impact neurodevelopmental and neuropsychiatric functions in the context of SUDs are not fully understood (Downey & Loftis, 2014).

Exosomes are small endosomal derived membrane microvesicles (40–140 nm in diameter) present in a variety of biofluids such as saliva, urine, and plasma, and they have a significant role in a series of fundamental biological processes including cellular activity regulation, intercellular communication, and the capacity to allow cells to exchange proteins, lipids, and genetic materials (Chen *et al*, 2019b; Huang *et al*, 2018; Zhang *et al*, 2018). The contents of exosomes are stable and the value of analyzing enriched exosomal DNA/RNA is greater than that of non-exosomal DNA/RNA, as the nucleotide sequence information is essentially diluted in the peripheral circulating blood that usually results in an increased signal-to-noise ratio (Cheng *et al*, 2015; Ramirez *et al*, 2018). As one of the most important contents in exosomes, miRNA are non-coding RNA species that can bind to complementary sites of the 3′ untranslated region of their mRNA targets resulting in the downregulation of gene expression. Notably, exosomes can migrate across the blood-brain barrier (BBB) (Chen *et al*, 2016; Saeedi *et al*, 2019) and their contents can be released into the extracellular environment by binding to RNA-binding proteins or through secretion in cell-derived plasma microvesicles. As a result, dysregulated exosomal miRNA expression profiling in peripheral circulation may therefore reflect physiological state and have diagnostic potential for neurological and neuropsychiatric disorders. For example, a novel 16-miRNA signature has been identified and including established risk factors (e.g., age, sex, and apolipoprotein □4 allele status) to a panel of dysregulated exosomal miRNA would result in a sensitivity and specificity of 87% and 77%, respectively, for predicting Alzheimer’s disease (Cheng *et al*., 2015). In addition, studies have shown that exosomes are involved in the transmission and release of α-synuclein, which are correlated with the severity of cognitive impairment in patients with Parkinson’s disease and dementia (Stuendl *et al*, 2016). Therefore, investigating exosomes may identify valuable biomarkers with diagnostic potential and offer the opportunity to uncover the regulatory role of peripheral circulating exosomes in substance dependence and withdrawal.

Although high-throughput sequencing has been attempted to explore the miRNA content in exosomes from patients with SUDs (Chen *et al*, 2021; Wang *et al*, 2019), the underlying molecular mechanisms of exosomal miRNAs in the regulation of withdrawal symptoms remains to be fully elucidated. In the present study, we aimed to profile neurotransmitter and exosomal miRNAs in plasma to identify a subset of neurotransmitter and exosomal miRNA biomarkers in a large group of patients with SUDs undergoing withdrawal. We subsequently explored the molecular function and possible mechanism of identified signature miRNA using an *in vitro* model of human induced pluripotent stem cells (iPSCs) derived neurons. Taken together, the analyses of neurotransmitter and miRNA content in peripheral circulating exosomes may strengthen our knowledge and provide valuable information contributing to the development of diagnosis and therapeutic interventions, and a mechanistic view of disease phenotypes during substance dependence and withdrawal.

## Materials and Methods

### Ethics statement and clinical sample collection

In this cross-sectional study, a total of 120 male patients with SUDs were recruited from a joint program for drug detoxification and rehabilitation in the First Affiliated Hospital of Kunming Medical University and the Kunming Drug Rehabilitation Center from January 2018 to October 2019. Twenty age-matched healthy male volunteers were recruited from the Physical Examination Center in the First Affiliated Hospital of Kunming Medical University. Prior to inclusion in the study, general demographic data and complete medical, psychiatric, and substance abuse histories for all participants were collected for all participants. Written informed consent for participation was provided by all individuals. The study was approved by the First Affiliated Hospital of Kunming Medical University Ethics Committee (2018-L-42). Each patient was diagnosed as having opioid (e.g., heroin) or amphetamine (e.g., methamphetamine) dependence based on the Diagnostic and Statistical Manual of Mental Disorders, 5th edition (DSM-5) criteria provided by the American Psychiatric Association. Individuals were excluded if they fulfilled any of the following conditions: any other DSM-5 axis I or II disorders (other than amphetamine/opioid dependence); HIV antibodies; neurological disorders or serious medical conditions; polysubstance use. Of the total 140 participants recruited, there were 60 heroin-dependent patients (HDPs, 20:20:20 at the 7-day, 3-month, and 12-month withdrawal stage), 60 methamphetamine-dependent patients (MDPs, 20:20:20 at the 7-day, 3-month, and 12-month withdrawal stage), and 20 healthy controls (HCs). An initial 10 participants per each group were used as the discovery cohort, and the subsequent 10 participants per each group were used as the validation cohort (**Figure 1A**). Peripheral blood samples were collected in 5 ml EDTA□2Na vacuum tubes between 8:00 and 10:00 AM in order to minimize a possible rhythm variance bias and the clinical data were obtained at the same time. Within 1 hour of blood collection, plasma was separated out by centrifugation and was immediately aliquoted and frozen in liquid nitrogen. The stored plasma samples were transported and packaged on dry ice.

**Figure 1:**
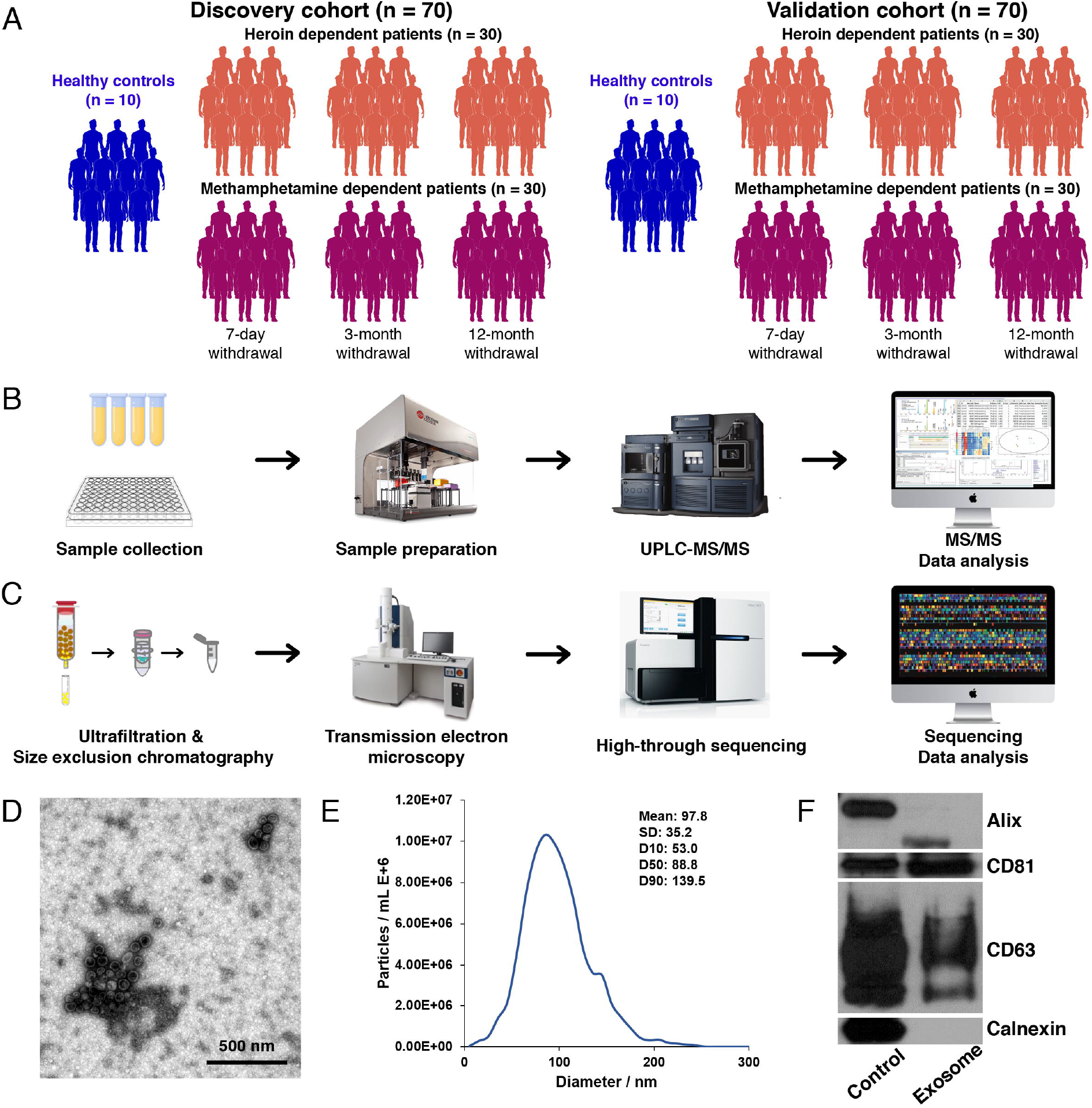
Pipeline showing the experimental design in this study. (A-C) An overview of the experimental design. (D-E) Characterisation of exosomes derived from the plasma of Substance use disorder (SUD) patients and controls; (D) The shape and structure of plasma exosomes isolated by ultracentrifugation under transmission electron microscopy (TEM); (E) The size of exosomes was analysed by nanoparticle tracking analysis (NTA); (F) Western blots of exosomal membrane markers, including Alix, CD63, and CD81. The exosomes isolated from each participant underwent both TEM, NTA and Western blotting to confirm successful enrichment and ensure limited contamination with cellular debris prior to small RNA deep sequencing.

### Scales

The Hamilton-Anxiety scale (HAM-A) is a semi-structured questionnaire administered by the interviewer, who assesses 14 clusters of questions, each concerning a series of symptoms. Seven elements examine psychological stress and seven examine physical stress with a satisfactory inter-rater reliability (Hamilton, 1959). The Hamilton-Depression Scale (HAM-D) is a semi-structured questionnaire administered by the interviewer, who assesses 21 clusters of questions, measuring depression symptoms (Hamilton, 1960).

### Sample preparation and instrumentation

Samples were thawed on ice-bath and 25 μl of each plasma sample was transferred to a 96-well plate. Then the plate was automated by Biomek 4000 workstation (Beckman Coulter, Inc., Brea, USA) following preprogrammed protocol. Briefly, 100 μl ice cold methanol with partial internal standards was automatically added to each sample and vortexed vigorously for 5 min. The plate was centrifuged at 4000g for 30 min (Allegra X-15R, Beckman Coulter, Inc., Indianapolis, USA). Next, 30 μl of supernatant was transferred to a clean 96-well plate, and 20 μl of freshly prepared derivative reagents was added to each well. The plate was sealed, and the derivatization was carried out at 30°C for 60 min. After derivatization, 350 μl of ice-cold 50% methanol solution was added to dilute the sample. Then the plate was stored at - 20°C for 20 min and followed by 4000g centrifugation at 4°C for 30 min. An ultra-performance liquid chromatography coupled to tandem mass spectrometry (UPLC-MS/MS) system (ACQUITY UPLC-Xevo TQ-S, Waters Corp., Milford, USA) was used to quantitate the metabolite in this project (**Figure 1B**).

### Sample processing

The rapid turnover of many intracellular metabolites makes immediate metabolism quenching necessary. The extraction solvents were stored in -20°C freezer overnight and added to the samples immediately after the samples were thawed. An ice-salt bath was used to keep samples at a low temperature to minimize degradation during sample preparation. All the prepared samples were analyzed within 48 hours after extraction and derivatization. A comprehensive set of rigorous quality control/assurance (QC/QA) procedures was employed to ensure a consistently high quality of analytical results, throughout controlling every single step from sample receipt at laboratory to final deliverables. The goal of QC/QA was to provide the reliable data for biomarker discovery study. To achieve this, three types of quality control samples (test mixtures, internal standards, and pooled biological samples) were used in the metabolomics platform. In addition to the quality controls, conditioning samples, and solvent blank samples were also used for obtaining optimal instrument performance. Each sample was accessioned into Metabo-Profile LIMS system and was assigned by the LIMS a unique identifier, which was used to track all sample handling, tasks, results and other conditions. The samples and aliquots were barcoded and analyzed by the LIMS system.

### Metabonomic data analysis

Data were analyzed using a XploreMET v3.0 system (Metabo-Profile, Shanghai, China). Partial least squares projection to latent structures and discriminant analysis (PLS-DA) were used to process the acquired GC–TOF/MS data. Principal component analysis (PCA) and partial least square discriminant analysis (PLS-DA) were performed for both positive and negative models after log transformation and pareto scaling. The variable importance in the projection value of each variable in the PLS-DA model was calculated to indicate its contribution to the classification. The significance of each metabolite with the VIP value >1 was assessed by Student’s *t*-test at univariate level, and the results were adjusted for multiple testing using the Benjamini–Hochberg procedure with the critical false discovery rate (FDR) set to 0.05.

### Enrichment of plasma derived exosomes from peripheral blood

Fresh whole blood samples from recruited patients or HCs were collected into the EDTA□2Na vacuum tubes and the blood was centrifuged at 1500 g for 15 min. The plasma was transferred into a new tube and centrifuged at 20,000 g at 4 °C for 15 min. The supernatant was collected and kept at −80 °C until use. The exosomes were isolated by size exclusion chromatography method as described previously with minor modifications (Boing *et al*, 2014). In brief, 2 ml of 0.8 μm-filtered plasma sample was purified using Exosupur® columns (Echobiotech, China). The exosome samples were eluted with PBS and a total of 2 mL eluate fractions were collected according to the manufacturer’s instruction. Fractions were concentrated to 200 μl by 100 kDa molecular weight cut-off Amicon® Ultra spin filters (Merck, Germany).

### Transmission electron microscopy and nanoparticle tracking analysis

For transmission electron microscopy analysis (Varga *et al*, 2014), 10 μl of exosomal suspension was loaded on a continuous carbon grid, and incubated for 10 min at room temperature, followed by distilled water washing. After being absorbed excessively, each sample was negatively stained with 10 μl of 2% (w/v) uranyl acetate solution at room temperature for 1 min and sucked up residue by filter paper. Then, the samples were photographed at 80kv by a Hitachi H-7650 transmission electron microscope (Hitachi, Tokyo, Japan). Particle size and concentration of plasma-derived EVs were analyzed by nanoparticle tracking analysis (NTA) on a Nano Sight NS300 instrument (Malvern Panalytical, Malvern, UK) following the manufacturer’s protocol (Varga *et al*., 2014). In brief, the samples were diluted to 1×10^7^ ∼ 1×10^9^ particles/ml in PBS, and slowly injected into the sample pool and detected in flow mode. These flow measurements consisted of three measurements of 60 s. The captured data were analyzed using NTA 3.2 software.

### Library preparation and small RNA sequencing

Total RNAs from exosomes were isolated using the miRNeasy® Mini kit (Qiagen, MD, USA) according to the manufacturer’s protocol. Sequencing libraries were generated using the QIAseq miRNA Library Kit (Qiagen, MD, USA), according to the manufacturer’s instructions. 500 ng of exosome derived total RNA for each sample was used as input for library preparation. The RNA samples were barcoded by ligation with unique adaptor sequences to allow pooling of samples. The elution containing pooled DNA library was further processed for cluster generation and sequencing using a NovaSeq 6000 platform.

### Real-time PCR analysis

To validate the miRNAs identified in RNA sequencing data, we performed qPCR analysis of selected miRNA targets. Total RNA extraction from the serum exosomes was performed with the miRNeasy® Mini kit (Qiagen, MD, USA) according to the manufacturer’s instructions. The concentration and quality of the RNA was determined using the RNA Nano 6000 Assay Kit of the Agilent Bioanalyzer 2100 System (Agilent Technologies, CA, USA). Single strand cDNA was synthesized with the PrimeScript™ RT reagent Kit (Takara, Dalian, China). The RT-PCRs were conducted on an ABI Prism 7500 HT using the TaqMan Fast Advanced Master Mix. Real-time PCR amplification was performed in triplicate and a negative control was included for each primer. miRNA levels were normalized to average Ct of internal control has-miR-106a-5p in the confirmation phase.

### Analysis of miRNA-predicted target genes and network visualization

The miRNet (http://www.mirnet.ca) was used to predict mRNAs targets, conduct functional enrichment analysis and generate network visualization (Fan *et al*, 2016). The miRNA-target interaction data were based on 10 well-annotated databases including miRTarBase, TarBase, miRecords, SM2miR, Pharmaco-miR, miR2Disease, PhenomiR, StarBase, EpimiR, miRDB, in which the datasets were merged on the same targets for each species, removed the duplicate entries, updated the literature references and stored them into SQLite database for fast retrieval. The conventional hypergeometric tests were implemented in miRNet for functional annotations-based Disease, GO, or KEGG pathway analysis.

### Stem cell culture and neurite outgrowth assay

Human induced pluripotent stem cells (hiPSCs) were purchased from Guidon Pharmaceutics (Beijing, China) and cultured in mTeSR™ Plus media (STEMCELL Technologies) for feeder-free maintenance and expansion. For neurite outgrowth assay, 0.7 x 10^5^ iPSCs/well were first seeded in a 24-well plate and neuronal differentiation from hiPSCs was conducted according to the protocol previous published (Zhang *et al*, 2013). Ectopic expression of targeted miRNA (pEZX-MR04 or pEZX-MR04-hsa-744-5p) via lentiviral transfection (250μl media + 50μl lentivirus + 4μg/ml polybrene) for 24 hours was applied on day 4 of neuronal differentiation. Three, five and seven days after lentiviral induction of targeted miRNA expression and passage to a new dish, hiPSCs derived neurons were imaged at 20x magnification using CellInsight™ CX5 High Content Screening Platform (Thermo Scientific). Individual cell measurements of mean/median/maximum process length, total neurite outgrowth (sum of the length of all processes), number of processes, number of branches, cell body area, and mean outgrowth intensity were obtained using the HCS Studio™ 2.0 Cell Analysis Software (Thermo Scientific). Total RNA from cells was isolated using miRNeasy® Mini kit (Qiagen, MD, USA) according to the manufacturer’s instructions.

### DiR-labelled exosomes and in vivo biodistribution assay

Female C57BL/6 mice (aged 6 to 8 weeks old) were kept under standard laboratory conditions. Mice were housed in a temperature-controlled room with a 12-hour light/dark cycle. The University of Kunming Medical University Committee on the Use and Care of Animals approved all experiments. The filtered exosome of serum sample from healthy donors was incubated with 1 mM fluorescent lipophilic tracer DiR (1,1-dioctadecyl-3,3,3,3-tetramethylindotricarbocyanine iodide) (D12731, Invitrogen, Life Technologies) at room temperature for 15 minutes prior to exosome isolation. Lipopolysaccharide (LPS) or PBS treated C57BL/6 mice (*n* = 3/group) were used and freshly purified DiR-labeled exosomes were injected through the tail vein for intravenous injections. The biodistribution of labeled exosomes was examined using 2.0 x 10^10^ particles/gram body weight (p/g). The particle count was measured with NTA and the samples were diluted accordingly. For the analysis of DiR-exosome distribution, IVIS Spectrum (Perkin Elmer) was used. Either live (isoflurane sedated) mice were imaged, or the animals were sacrificed and then the organs harvested prior to analysis. For the perfusion experiment, the mice were sedated, and the vascular system was flushed by transcardial perfusion for 5 minutes. The left ventricle was infused with PBS (5 ml/min) and the right atrium was perforated. After perfusion, the organs were harvested and analyzed.

### Statistical analysis and bioinformatics analysis

All data were analyzed with R version 3.6.3. Kruskal–Wallis test or analysis of variance (ANOVA) test followed by a post-hoc test (Bonferroni’s t test) was used to test for differences in continuous variables. Comparison between groups was performed using a Student’s t test or a Chi-square analysis as appropriate. The Pearson correlation was used for correlation analyses. For miRNA sequencing, the raw fastq files generated from next-generation sequencing were preprocessed to remove adaptor sequences using the TagCleaner. Subsequently, known and novel miRNAs were detected within the preprocessed data using the miRDeep2 (Friedlander *et al*, 2012). Statistical analysis and significance were calculated using various tests and adjusted for multiple testing. Adjusted statistical significance was assessed as p < 0.05.

## Results

### Characteristics of the study population

A total of 120 patients with SUDs and 20 age-matched healthy participants (without SUDs) were recruited at the First Affiliated Hospital of Kunming Medical University. The characteristics of the study participants are presented in **Table 1**. There were no significant differences for any variables including age, BMI, substance dependence history, annual income, and education level between patients with SUDs and healthy participants in both discovery and validation cohorts (**Table 1**).

**Table 1:** Clinical and Demographic characteristics of study participants.

|  | Discovery cohort |  |  | Validation cohort |  |  | Discovery.p | Validation.p | Discovery.p | Validation.p | Discovery.p | Validation.p |
| --- | --- | --- | --- | --- | --- | --- | --- | --- | --- | --- | --- | --- |
|  | Control<br>(n=10) | Heroin<br>(n=30) | Methamphetamine<br>(n=30) | Control<br>(n=10) | Heroin<br>(n=30) | Methamphetamine<br>(n=30) | Control vs Heroin |  | Control vs<br>Methamphetamine |  | Heroin vs<br>Methamphetamine |  |
| Age (year) | 36.71±7.32 | 37.25±7.59 | 36.58±7.72 | 36.03±9.05 | 36.84±8.27 | 36.28±8.42 | 0.7667 | 0.7906 | 0.9875 | 0.9805 | 0.7117 | 0.6467 |
| BMI | 21.36±1.68 | 22.05±1.58 | 21.56±1.47 | 22.01±1.6 | 21.38±1.89 | 21.58±1.28 | 0.3903 | 0.3408 | 0.9378 | 0.5739 | 0.3042 | 0.5106 |
| History (year) | NA | 7.8±2.52 | 7.46±2.78 | NA | 6.79±2.97 | 7.06±2.51 | NA | NA | NA | NA | 0.2976 | 0.6467 |
| Education* | 1/6/2/1 | 9/12/7/2 | 9/11/9/1 | 1/4/4/1 | 6/10/9/5 | 7/9/9/5 | 0.5682 | 0.8801 | 0.3203 | 0.8460 | 0.9184 | 1.0000 |
| Income** | 0/3/4/3/0 | 5/12/7/4/2 | 5/11/5/7/2 | 0/4/4/2/0 | 5/12/8/4/1 | 5/11/5/7/2 | 0.4330 | 0.7015 | 0.4111 | 0.4595 | 0.9083 | 0.7684 |
| HAM-A | 4.9±2.42 | 12.25±5.86 | 11.3±5.1 | 5.4±2.17 | 14.35±3.66 | 13.55±4.58 | 0.0005 | 0.0000 | 0.0014 | 0.0001 | 0.7245 | 0.6935 |
| HAM-D | 8.2±3.82 | 12.95±5.89 | 13.45±5.99 | 8.8±3.05 | 13.15±6.72 | 13.8±5.68 | 0.0134 | 0.1708 | 0.0170 | 0.0273 | 0.7140 | 0.8283 |

Data are mean ± SD. Group differences are evaluated with Kruskal’s test for continuous variables and fisher’s test for categorical variables. \* Education levels: illiteracy/primary school/middle school/college; \*\* Income levels: monthly 0~1000¥/1000~3000¥/3000~5000¥/5000~10000¥/10000+¥. HAM-A: Hamilton Rating Scale for Anxiety; HAM-D: Hamilton Depression Rating Scale.

Substance use, depression, and anxiety frequently co-occur (Lai *et al*, 2015). To better understand whether the use of heroin or methamphetamine influences mood (specifically, anxiety and depression), the HAM-A and HAM-D questionnaires were administered by experienced interviewers. As expected, the HAM-A total scores were significantly increased in the HDPs (*p* = 0.0005) and the MDPs (*p* = 0.0014) when compared to HCs in the discovery cohort **(Table 1)**. Significant increases relative to HCs were observed in both HDPs and MDPs at the 3-month and 12-month stages after the initiation of withdrawal, respectively (**Table S1**). The overall differences in HAM-A for HDPs and MDPs relative to HCs, as well as the differences in the subgroups at the 3-month and 12-month withdrawal stages, were validated (**Table S2**). In contrast, the significant differences in HAM-D scores observed in the HDPs (*p* = 0.0134) in the discovery cohort did not persist in the validation cohort. In the subsequent subgroup analysis, the significant increases in HAM-D scores were only validated in the subgroup of MDPs at the 3-month withdrawal stage, but not in any of the subgroups for HDPs (**Tables S1 & S2**).

### Dynamic changes in peripheral neurotransmitters during substance withdrawal

In order to reveal peripheral neurotransmitter changes in the patients with SUDs, we analyzed the classic neurotransmitters and their metabolites, including γ-aminobutyric acid (GABA), glutamate, serotonin, tryptophan, kynurenine, choline, etc, through UPLC-MS/MS (**Table S3 and Figure 1B**). Plasma samples from the 70 participants in the discovery cohort were analyzed first for the 14 neurotransmitter metabolites. Of these, seven fell below the lower level of quantitation in > 20% of samples and were excluded from analysis. Detailed results for the seven neurotransmitters are presented in **Table 2**. Our analysis revealed that three of the seven neurotransmitters were associated with significant changes. Two neurotransmitters, GABA and choline, were significantly decreased in both HDPs (*fdr*.*p*.GABA = 0.0026, *fdr*.*p*.choline = 0.026) and MDPs (*fdr*.*p*.GABA = 0.0009, *fdr*.*p*.choline = 0.026) relative to HCs, respectively. In contrast, serotonin was significantly increased in HDPs (*fdr*.*p*.serotonin = 0.0001) and MDPs (*fdr*.*p*.serotonin = 0.0013) relative to HCs (**Table 2 and Figure S1**). These significant differences in plasma GABA, choline, and serotonin were further validated in the independent cohort (**Table S4**). Interestingly, there were no significant differences in the tested neurotransmitters between HDPs and MDPs (**Table 2 & S4**).

**Table 2.** Detailed information of the 7 neurotransmitters isolated from peripheral blood plasma of HC, HDPs and MDPs from the discovery cohort.

| Metabolites | *mean* | *mean* | *mean* | *p* | *fdr.p* | *p* | *fdr.p* | *p* | *fdr.p* |
| --- | --- | --- | --- | --- | --- | --- | --- | --- | --- |
|  | Control<br>(n=10) | Heroin<br>(n=30) | Methamphetamine<br>(n=30) | Control vs Heroin |  | Control vs Methamphetamine |  | Heroin vs Methamphetamine |  |
| Choline | 3741.63± 1408.82 | 2335.26±972.98 | 2395.46±1192.87 | **0.0127\*** | **0.0260\*** | **0.0173\*** | **0.0260\*** | 0.8352 | 0.8352 |
| GABA | 30.53 ± 7.51 | 19.73±9.70 | 17.81±7.20 | **0.0017\*** | **0.0026\*** | **0.0003\*** | **0.0009\*** | 0.4028 | 0.4028 |
| Glutamic acid | 43307.29 ± 16338.27 | 31349.56±10167.58 | 36624.70±15934.20 | 0.0516 | 0.1549 | 0.1744 | 0.2617 | 0.3057 | 0.3057 |
| Glutamine | 8414.58 ± 2173.78 | 7771.86±1693.76 | 8089.17±1192.05 | 0.4119 | 0.6012 | 0.6012 | 0.6012 | 0.4189 | 0.6012 |
| Kynurenine | 1406.28 ± 328.97 | 1397.10±322.75 | 1464.27±239.13 | 0.7567 | 0.7567 | 0.2195 | 0.5664 | 0.3776 | 0.5664 |
| Serotonin | 44.7 1± 44.32 | 240.15±266.50 | 251.18±388.72 | **4.8255E-5\*** | **0.0001\*** | **0.0009\*** | **0.0013\*** | 0.5955 | 0.5955 |
| Tryptophan | 24923.06 ± 5255.84 | 21501.42±4011.26 | 22417.54±4106.69 | 0.0838 | 0.2514 | 0.1938 | 0.2907 | 0.3979 | 0.3979 |

Next, to investigate potentially differential neurotransmitters in SUDs across the various withdrawal stages, we performed additional subgroup analyses. Notably, significant increases in serotonin were observed in all three subgroups of HDPs and two subgroups of MDPs when compared to HCs, but not observed in the subgroup of MDPs at the 12-month withdrawal stage (**Table 3**). In contrast, the significant decrease in GABA persisted in all three subgroups of MDPs relative to HCs but was only found in the HDPs at the 3-month withdrawal stage. Moreover, the significant decreases in choline were only observed in the subgroups of HDPs and MDPs at the 3-month withdrawal stage (**Table 3**). To validate these findings, the plasma neurotransmitter profiles across the three withdrawal stages were further tested in an independent cohort. Compared to HCs, the significant differences in serotonin at the 7-day withdrawal stage, as well as in GABA, choline, and serotonin at the 3-month withdrawal stage were validated in both HDPs and MDPs, respectively (**Table S5**).

**Table 3.**
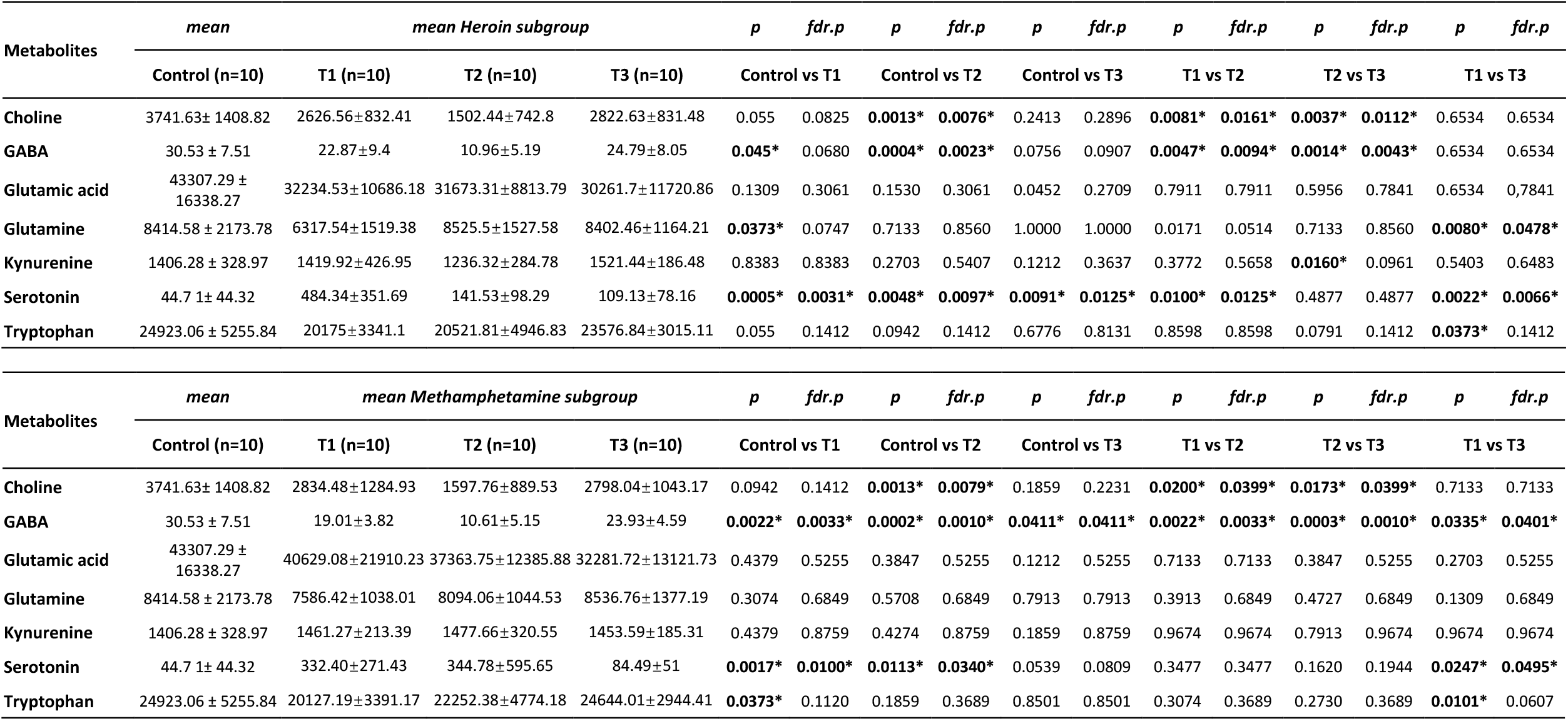
Detailed information of the 7 neurotransmitters isolated from peripheral blood plasma in HDPs and MDPs across three withdrawal stages compared to HC.

### Associations of neurotransmitters and psychiatric comorbidities in SUD patients

To further determine the associations between neurotransmitters and psychiatric comorbidities in SUDs, Pearson correlations between neurotransmitters (GABA, choline, and serotonin) and HAM-A/D scales were conducted. The plasma GABA in HDPs (R = -0.40, *p* = 0.01) and MDPs (R = -0.35, *p* = 0.03) was significantly correlated with HAM-A scores. However, the significant correlation between GABA and HAM-D scores was only observed in MDPs (R = -0.43, *p* = 0.0052) but not HDPs (R = -0.20, *p* = 0.21). In addition, significant correlations were found between plasma serotonin and HAM-A score in both HDPs (R = 0.44, *p* = 0.0054) and MDPs (R = 0.40, *p* = 0.0012). Nevertheless, no significant correlation was observed between serotonin and the HAM-D score, or between choline and the HAM-A/D scores (**Figure S2**). In summary, these results indicate some strong correlations between specific plasma neurotransmitters and withdrawal symptoms and suggest that the dynamic changes in peripheral GABA and serotonin may play a key role in influencing moods during withdrawal.

### Isolation and characterization of plasma exosomes

Despite research efforts in the field since the discovery of exosomes in peripheral circulation (Raposo & Stoorvogel, 2013), the role of exosomes in the pathophysiology of psychiatric disorders, and especially SUDs, remains poorly understood. In the present study, plasma exosomes were isolated from patients with SUDs and age-matched HCs by size exclusion chromatography in the discovery cohort, and their morphology was verified by transmission electron microscopy (TEM) and nanoparticle tracking analysis (NTA), as shown in **Figure 1D & E**. These analyses revealed an average particle diameter of 40–140 nm, and there was no significant difference in the exosome size distribution between the patients with SUDs and HCs (data not shown). In addition, the exosomes were verified by detecting the expression of exosomal surface markers CD81 and CD63 (Ricklefs *et al*, 2019) using western blotting (**Figure 1F**), which confirmed the successful exaction of exosomes from plasma samples.

### Differential expression of exosomal miRNA between SUD patients and healthy controls

To gain insight into the transcriptional dynamics of exosomes, small RNA sequencing was applied to the total RNA derived from plasma exosomes in all 70 participants in the discovery cohort. Sequencing quality statistics are shown in **Table S6**. Top RNA species detected by sequencing included miRNA, piRNA, tRNA, and snoRNA, and the percentage of reads assigned to each species and the number of mapped miRNAs did not significantly differ between samples (**Figure S3**). Length distribution analysis of the miRNAs showed that 20–24 nucleotides (nt) was the most frequent read length, with the peak at 22 nt (**Table S7**), indicating that mature miRNAs were suitably enriched during the sequencing library preparation process. To focus on the miRNA ubiquitously abundant in plasma and suitable as biomarker candidates, we selected the miRNA candidates with an absolute fold change in expression of ≥ 1.5, a nominal FDR value of ≤ 0.05, and the expression (RPKM ≥ 0) across ≥ 50% of samples. This yielded 43 and 26 miRNAs with significant changes in HDPs and MDPs relative to HCs, respectively (**Figure 2A-D, Table S8 & S9**). The Venn diagram displays a list of 20 shared differentially expressed miRNAs (DE-miRNAs) in HDPs and MDPs when compared to HCs (**Figure 2E, Table 4**), suggesting there might be a variety of shared, exosomal miRNA-associated, substance-related functional abnormalities between HDPs and MDPs.

**Table 4.**
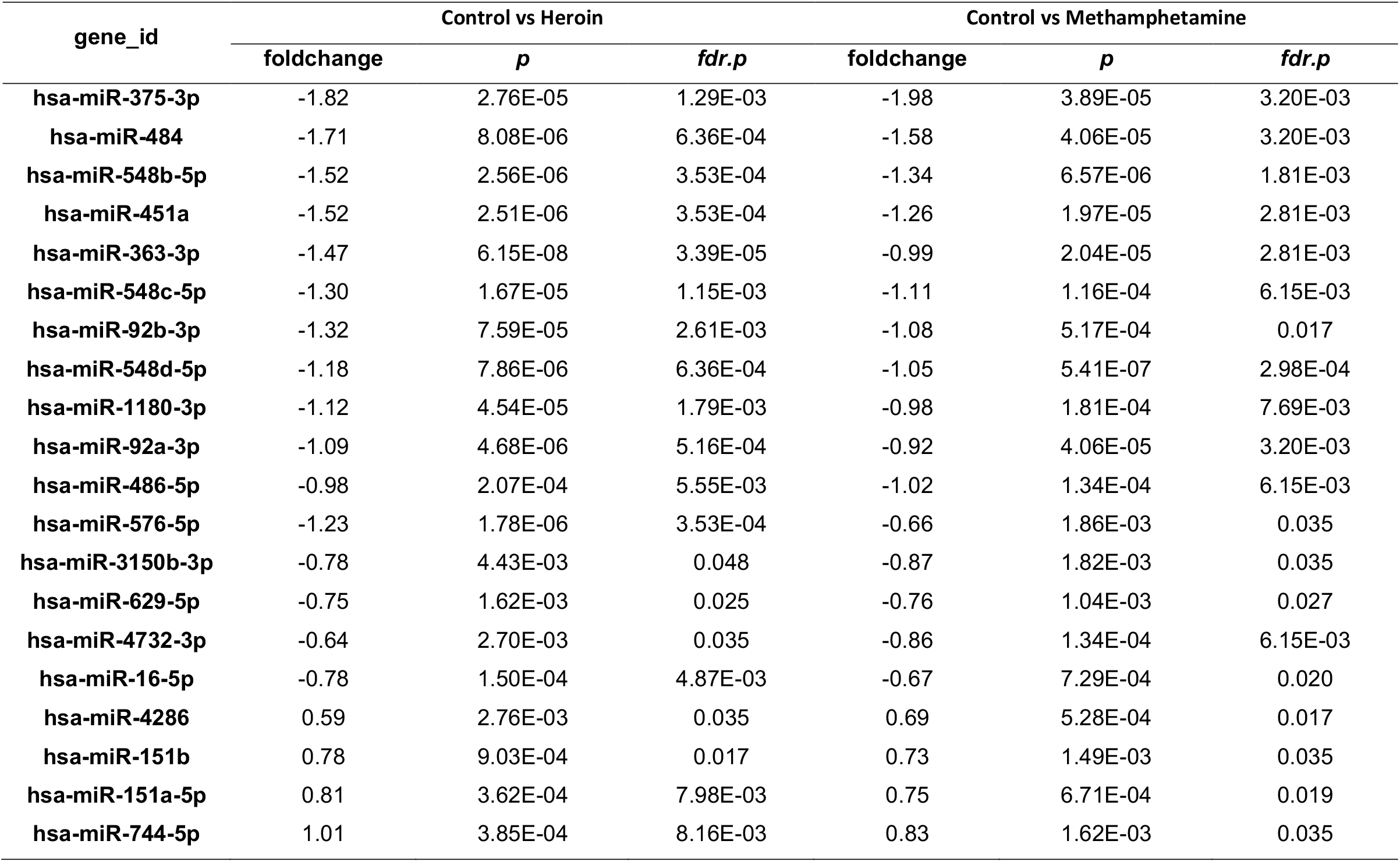
Significant miRNAs that change in exosomes derived from both HDPs and MDPs.

| gene_id | Control vs Heroin |  |  | Control vs Methamphetamine |  |  |
| --- | --- | --- | --- | --- | --- | --- |
|  | foldchange | *p* | *fdr.p* | foldchange | *p* | *fdr.p* |
| hsa-miR-375-3p | -1.82 | 2.76E-05 | 1.29E-03 | -1.98 | 3.89E-05 | 3.20E-03 |
| hsa-miR-484 | -1.71 | 8.08E-06 | 6.36E-04 | -1.58 | 4.06E-05 | 3.20E-03 |
| hsa-miR-548b-5p | -1.52 | 2.56E-06 | 3.53E-04 | -1.34 | 6.57E-06 | 1.81E-03 |
| hsa-miR-451a | -1.52 | 2.51E-06 | 3.53E-04 | -1.26 | 1.97E-05 | 2.81E-03 |
| hsa-miR-363-3p | -1.47 | 6.15E-08 | 3.39E-05 | -0.99 | 2.04E-05 | 2.81E-03 |
| hsa-miR-548c-5p | -1.30 | 1.67E-05 | 1.15E-03 | -1.11 | 1.16E-04 | 6.15E-03 |
| hsa-miR-92b-3p | -1.32 | 7.59E-05 | 2.61E-03 | -1.08 | 5.17E-04 | 0.017 |
| hsa-miR-548d-5p | -1.18 | 7.86E-06 | 6.36E-04 | -1.05 | 5.41E-07 | 2.98E-04 |
| hsa-miR-1180-3p | -1.12 | 4.54E-05 | 1.79E-03 | -0.98 | 1.81E-04 | 7.69E-03 |
| hsa-miR-92a-3p | -1.09 | 4.68E-06 | 5.16E-04 | -0.92 | 4.06E-05 | 3.20E-03 |
| hsa-miR-486-5p | -0.98 | 2.07E-04 | 5.55E-03 | -1.02 | 1.34E-04 | 6.15E-03 |
| hsa-miR-576-5p | -1.23 | 1.78E-06 | 3.53E-04 | -0.66 | 1.86E-03 | 0.035 |
| hsa-miR-3150b-3p | -0.78 | 4.43E-03 | 0.048 | -0.87 | 1.82E-03 | 0.035 |
| hsa-miR-629-5p | -0.75 | 1.62E-03 | 0.025 | -0.76 | 1.04E-03 | 0.027 |
| hsa-miR-4732-3p | -0.64 | 2.70E-03 | 0.035 | -0.86 | 1.34E-04 | 6.15E-03 |
| hsa-miR-16-5p | -0.78 | 1.50E-04 | 4.87E-03 | -0.67 | 7.29E-04 | 0.020 |
| hsa-miR-4286 | 0.59 | 2.76E-03 | 0.035 | 0.69 | 5.28E-04 | 0.017 |
| hsa-miR-151b | 0.78 | 9.03E-04 | 0.017 | 0.73 | 1.49E-03 | 0.035 |
| hsa-miR-151a-5p | 0.81 | 3.62E-04 | 7.98E-03 | 0.75 | 6.71E-04 | 0.019 |
| hsa-miR-744-5p | 1.01 | 3.85E-04 | 8.16E-03 | 0.83 | 1.62E-03 | 0.035 |

**Figure 2:**
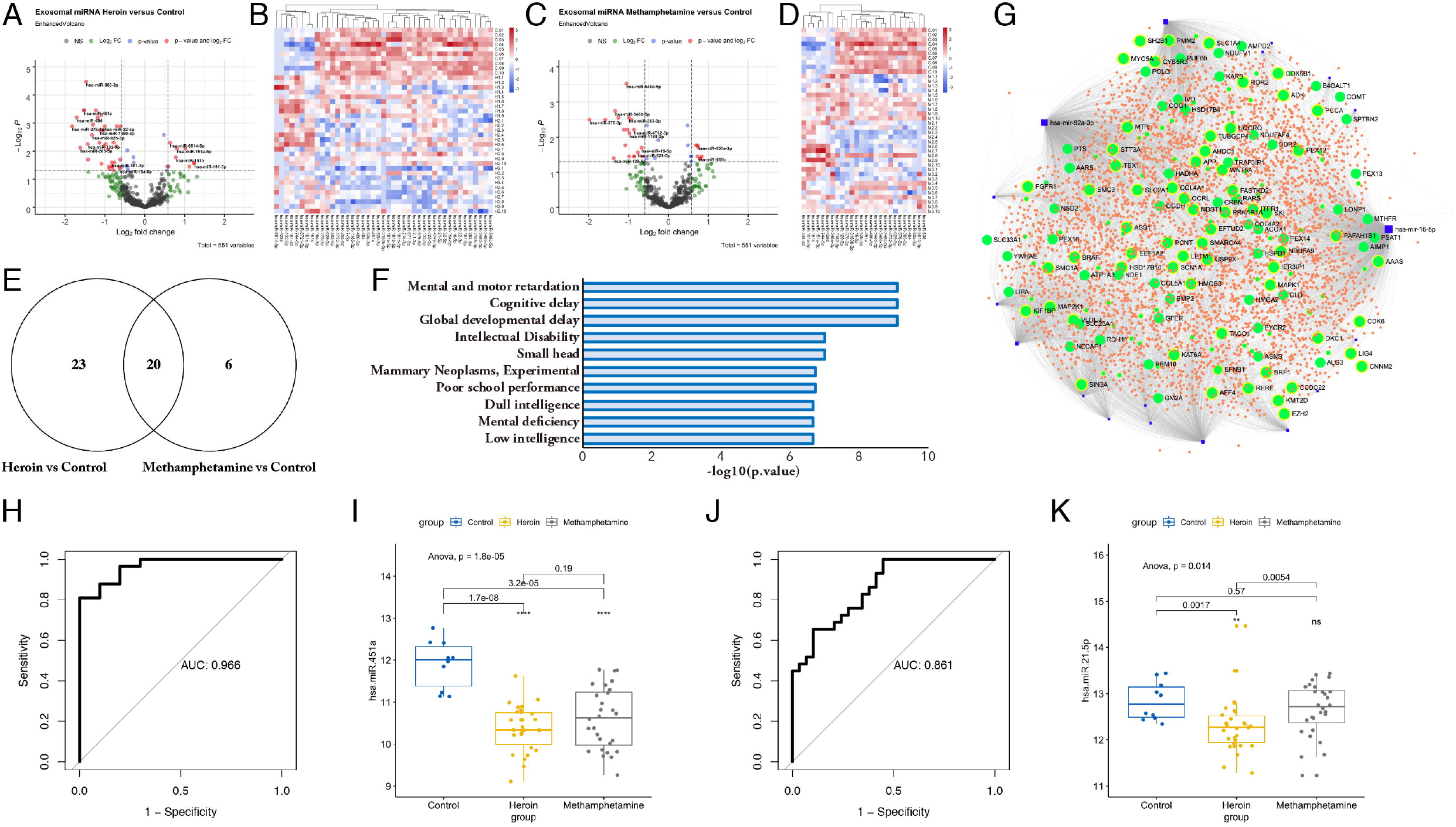
Distinct exosomal miRNAs in HC, HDPs and MDPs. Small RNA sequencing was performed to reveal the small RNA content in plasma samples collected from HC and patients with SUDs. (A-D) Statistically significant (Fold change ≤ −1.5 and ≥ 1.5, *p*.*fdr* ≤ 0.05) tRNA fragments were detected in (A, B) HDPs vs HC and (C, D) MDPs vs HC. (E) Venn diagram showing the number of the DE-miRNAs identified in HDPs and MDPs compared to HC. (F, G) Predicted functional pathways and biological functions (F) associated with mRNA targets of the 20 overlapped DE-miRNAs and (G) miRNA-mRNA interacting network. (H) ROC AUC for the diagnosis of patients with SUDs compared to HC. (I) Expression of miR-451a in HC, HDPs and MDPs. (J) ROC AUC for the diagnosis of MDPs compared to HDPs. (K) Expression of miR-21 in HC, HDPs and MDPs. The range of expression values in the groups is indicated by the error bars.

### Pathway analysis and gene target and gene ontology prediction

Network visualization and functional enrichment analysis is a powerful combination in delivering important biological insights. Using miRNet, a network-based visual exploration and functional interpretation platform (Chang *et al*, 2020), we identified predicted functional pathways associated with mRNA targets of the 20 shared DE-miRNAs and thereafter generated an miRNA-mRNA interacting network (**Figure 2E-G**). Top associated pathway/disease hits included mental and motor retardation (*p* = 5.2e-24), cognitive delay (*p* = 4.2e-23), global developmental delay (*p* = 3.8e-22), with a strong overall theme of developmental or intellectual abnormality (**Figure 2F-G**). Next, the differential metabolites and miRNAs were combined and used to perform ROC analysis to identify an optimal metabolite and/or exosomal miRNA biomarker panel in plasma for SUDs. Strikingly, from the ROC curves, the calculated sensitivities and specificities of the miRNA in diagnosing SUDs and distinguishing HDPs and MDPs were identified (**Figure 2H-K**). Notably, the analysis of hsa-mia-451a and hsa-mir-21a resulted in an AUC of 0.966 and 0.861, suggesting that these plasma exosomal miRNA, especially has-miR-451a and has-miR-21-5p, could serve as additional indicators for monitoring substance dependence and withdrawal. In order to verify the accuracy of miRNA-seq, hsa-miR-451a and hsa-miR-21-5p were selected as mentioned above for RT-qPCR analysis. The results indicated that the expression patterns of hsa-miR-451a and hsa-miR-21-5p were similar between small RNA-Seq and RT-qPCR (**Figure S4**).

### Identification of exosomal miRNA signatures across heroin and methamphetamine withdrawal

We further investigated the changes in exosomal miRNAs in SUDs across the three withdrawal stages. When small RNA-seq data were subjected to weighted gene co-expression network analysis (WGCNA) (Langfelder & Horvath, 2008), multiple gene-network modules were obtained. We decided to focus mainly on the modules that could represent the dynamic changes of miRNA expression throughout withdrawal stages. Analysis of the module-trait relationship identified six unique modules (blue, brown, magenta, purple, pink, and cyan) correlated with the type of substance use or the stage of withdrawal (*p* < 0.05). The six modules represent different miRNA expression alteration patterns (**Figure 3A-B**). Hub gene/miRNA network analysis of the modules revealed that a hierarchical organization of highly connected gene/miRNA in each module, through key controlling genes/miRNAs in the modular network could be identified. Notably, the blue module miRNAs (e.g., hsa-miR-140-3p) were mainly elevated at the 3-month withdrawal stage for both HDPs and MDPs (**Figure 3C-D**). The purple module consisting of miRNAs significantly decreased at all three withdrawal stages (e.g., hsa-miR-92a-3p and hsa-miR-451a, **Figure 3E-F**). Different from the miRNAs in the blue module which showed an increase at the 3-month withdrawal stage, the miRNAs in the brown module were significantly increased at both the 7-day and 3-month stages (e.g., hsa-miR-139-3p, **Figure 3G-H**). By contrast, the miRNAs in the pink module were significantly decreased at both the 3-month and 12-month stages (e.g., hsa-miR-375-3p, **Figure 3I-J**). Moreover, the miRNAs in modules magenta and cyan had other interesting but unique expression patterns across withdrawal stages (**Figure 3K-N**).

**Figure 3:**
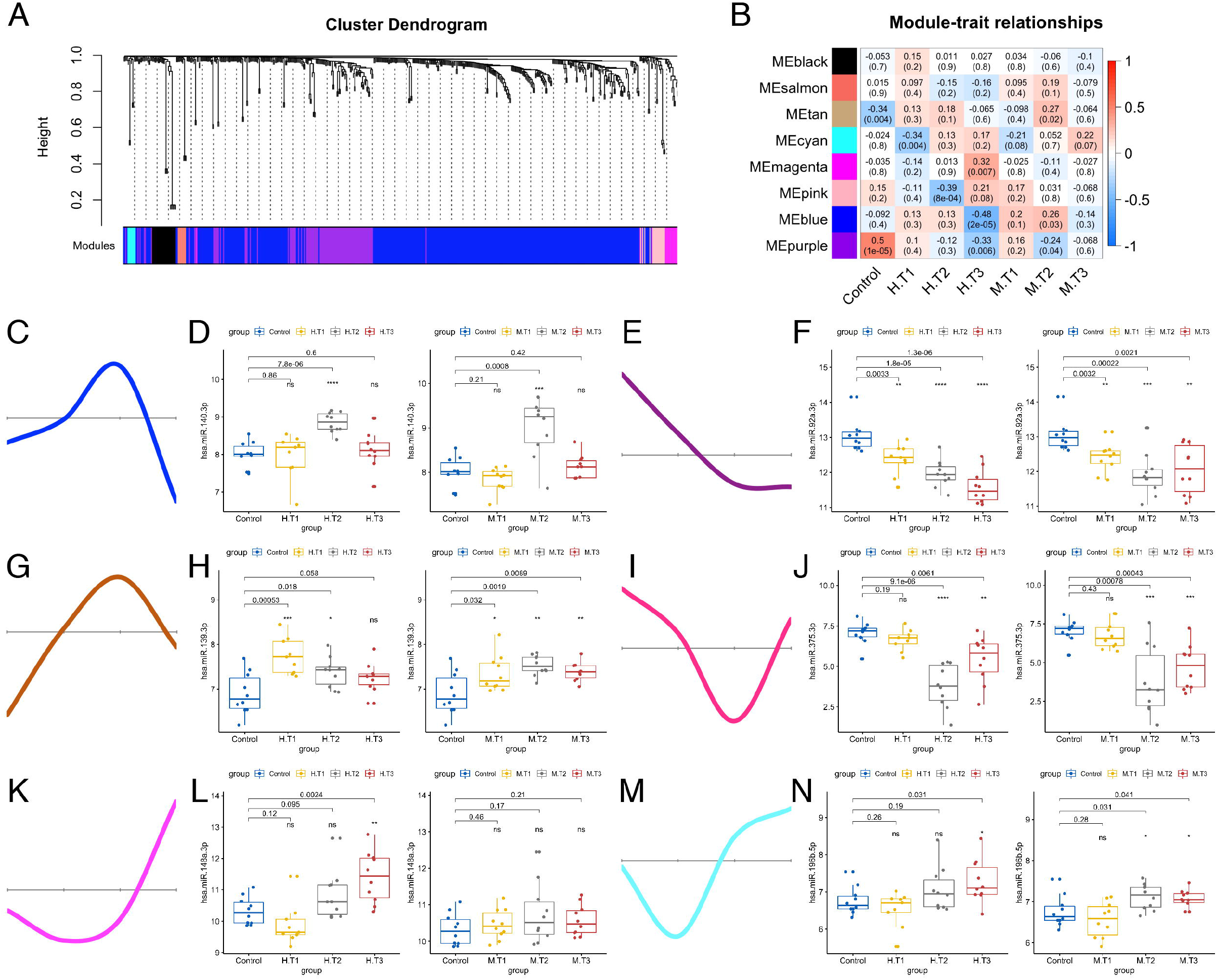
WGCNA of plasma exosome samples uncovered dynamic changes of miRNA signatures during substance withdrawal. (A) Hierarchical cluster dendrogram using WGCNA analyzing miRNA sequencing data. A total of 8 modules were identified after 0.25 threshold merging. (B) Module trait correlation analysis revealed that several key modules were significantly correlated with substance use and/or different withdrawal stages. (C, E, G, I, K, M) Eigengene expression of six selected modules across 4 groups of samples. Color code of the modules is preserved. (D, F, H, J, L, N) Expression patterns of key miRNAs representing six selected modules across subgroups of HDPs and MDPs.

We then sought to investigate the relationships between the neurotransmitters (GABA, choline, and serotonin) and the hub gene/miRNA in the blue and brown modules using Spearman’s correlations. One exciting finding was that hsa-miR-140-3p from the blue module showed strong negative correlations (*p* < 0.01) with both GABA and choline (**Figure S5A-B**). The hsa-miR-744-5p in the brown module was positively correlated with serotonin in HDPs (*p* = 4.9e-5) and MDPs (*p* = 0.032) (**Figure S5C**). Taken together, the WGCNA and the correlation analyses revealed a dynamic expression pattern of miRNA signatures during both heroin and methamphetamine withdrawal, and a range of neurotransmitter associated exosomal miRNAs in the peripheral blood plasma of patients with SUDs.

### Effects of hsa-miR-744-5p on neurite outgrowth in human iPSC derived neurons

Emerging studies have indicated an important role for specific miRNA in neural differentiation, proliferation, and maturation during embryonic development and/or disease progression using neurite outgrowth assays (Agostini *et al*, 2011; Le *et al*, 2009). To ascertain the neuronal function and molecular mechanism of hsa-miR-744-5p, we differentiated hiPSCs to neurons and infected them with lentivirus-miR-744-5p or scrambled control (**Figure 4A-B**). As illustrated in **Figure 4C**, across three replicate experiments, when iPSC derived neurons were treated with a scrambled control, there was no decrease in relative total outgrowth and cell viability. By contrast, overexpression of hsa-miR-744-5p was sufficient to cause significant decreases in neurite processes (**Figure 4D-F**). For further characterization, genome-wide gene expression changes upon miRNA transfection were studied using RNA-seq and comparative transcriptome analysis on hiPSCs derived neuronal culture with ectopic hsa-miR-744-5p expression versus scrambled control. One day after the lentivirus infection, a list of 46 genes were differentially expressed with a cut-off (fold change ≤ −1.5 or ≥ 1.5, *p* ≤ 0.05), in which the differentially expressed genes (DEGs) were mainly over-represented in axon development (*p* = 2.0e-3), reproductive process (*p* = 5.5e-3) and system development (*p* = 7.6e-3) (**Figure 4G-H**). Likewise, 556 DEGs with the same cut-off were identified on day 3 after the induction of hsa-miR-744-5p expression in hiPSCs derived neuronal culture and these DEGs were mainly enriched for cell cycle arrest (*p* = 1.1e-3), cell growth (*p* = 1.8e-3), and positive regulation of neuron apoptotic process (*p* = 2.4-3) (**Figure 4I-J**). RNA-seq data may partly demonstrate secondary effects on global mRNA expression, while gene expression changes are triggered by primary miRNA targets. Therefore, to discriminate between primary and secondary target genes, the differential expression results obtained from the RNA-seq analysis were compared to potential hsa-miR-744-5p targets predicted by bioinformatic algorithms. The overlap of statistically significant DEGs and bioinformatically predicted target genes revealed that hsa-miR-744-5p may play a significant role involving neural development and function regulating its primary miRNA targets PSENEN, FDX1L, VPS37D, SPTBN4, PPP5C, and FAM57B (**Figure 4K)**, although their mechanisms require further investigation. Overall, the neurite outgrowth assays and RNA-seq analyses reveal an important role of hsa-miR-744-5p in maintaining neuronal development and function.

**Figure 4:**
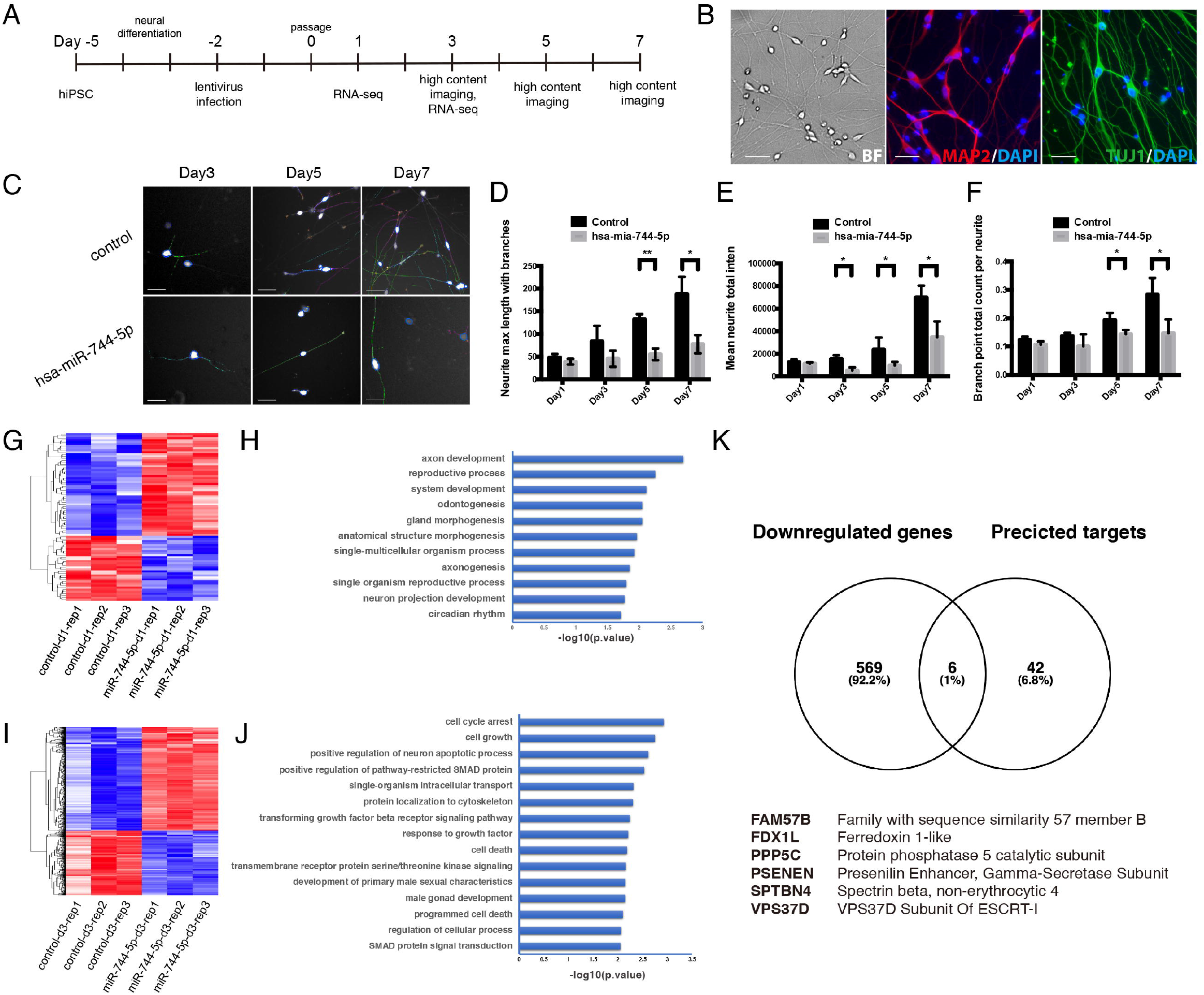
Effects of hsa-miR-744-5p on neurite outgrowth in human iPSC derived neurons. (A) Flow diagram of human iPSC derived neuron cell generation. (B) Representative differential interference contrast (DIC), MAP2 (Red) and TUJ1 (Green) fluorescence images illustrating the generation of human iPSC derived neurons cell. (C) Representative images illustrating the effects of hsa-miR-744-5p or control in neurite outgrowth assay on Day3, Day5 and Day7. (D-F) Statistics of neurite outgrowth assay after induction of hsa-miR-744-5p expression in human iPSC derived neurons. All data are presented as mean ± SD; n ≥ 3. p < 0.05, one-way ANOVA with Bonferroni post hoc analysis.

## Discussion

Herein, we report the dynamics of neurotransmitter and exosomal miRNA profiles in the peripheral circulation of 120 patients with SUDs and 20 healthy participants using UPLC-MS/MS and high-throughput sequencing technologies. After controlling for various factors (age, BMI, substance dependence history, annual income, and education), we successfully identified a set of critical miRNAs in circulating exosomes that were associated with neurotransmitters and the psychological comorbidities in these patients undergoing withdrawal. *In vitro* studies showed that increased hsa-miR-744-5p, one of key exosomal miRNAs, inhibited neurite outgrowth human iPSC derived neurons and may play a significant role in neural function by crossing BBB and regulating its primary gene targets.

Detoxification represents an important initial phase in the long-term recovery process by relieving withdrawal discomfort (Bart, 2012). To date, although a cascade of neurotransmitter and neuroendocrine abnormalities and neuronal damage are well-recognized consequences following long-term abuse of illegal substances, only limited molecular biomarkers have been identified for the associated symptoms (Volkow *et al*, 2015). Diagnosis is thus mainly left to the use of subjective questionnaires, particularly during the detoxification stage within the first 3 to 6 months (Stein *et al*, 2020; Timko *et al*, 2019). We observed that the plasma serotonin was significantly increased in patients with SUDs and anxiety (HAM-A) was significantly correlated with higher plasma serotonin concentration as compared with HCs, supporting the notion that changes in plasma serotonin seem to be more associated with the presence of psychiatric comorbidity than other neurotransmitters such as GABA and choline (Araos *et al*, 2019; Fava *et al*, 2018). While changes in the kynurenine pathway appear to be directly associated with a history of alcohol use disorder (AUD), altered tryptophan concentrations are associated with the presence of comorbid psychiatric disorders (Vidal *et al*, 2020). However, we did not observe a significant change in plasma kynurenine or tryptophan in individuals with SUDs, suggesting that AUD and SUDs might have different underlying mechanisms.

In the literature, emerging evidence has been documented for the importance of exosomes in the diagnosis, prognosis, and therapy of neurological and psychological disorders (van Niel *et al*, 2018). Exosomes carry abundant mRNA, DNA, miRNA, lncRNA, and other nucleic acid species, which provides strong support for the use of exosomes as biomarkers obtained via liquid biopsy (O’Brien *et al*, 2020). Specifically, the contents of exosomes are stable and can be delivered into recipient cells to exert their biological functions, indicating that the exosome-derived DNA/RNA could be used to account for the molecular regulation of diseases and reflect the conditional state/progression. Using small RNA sequencing, the present study uncovered insights into the dynamic expression pattern of miRNAs in the exosomes at various stages of substance withdrawal. First, we have identified a set of dysregulated miRNA signatures including hsa-mia-451a and hsa-mir-21a, with an AUC of 0.966 and 0.861, respectively, for predicting and distinguishing patients with HDPs and MDPs. Second, specific trends of change (i.e., increases, decreases, and combinations of both) of miRNA content in exosomes as a function of withdrawal stage were also identified. Although the underlying mechanism require further investigation, the possible functions of several dysregulated exosomal miRNAs have been proposed in other studies. For example, ethanol exposure increases hsa-miR-140-3p in extracellular vesicles, which may result in aberrant neural progenitor cell growth and maturation, explaining brain growth deficits associated with prenatal alcohol exposure (Keller *et al*, 2017; Tseng *et al*, 2019). We showed that the exosomal hsa-miR-140-3p is mainly elevated at the 3-month withdrawal stage in plasma for both HDPs and MDPs, and it was negatively correlated plasma GABA and choline, indicating that the plasma exosomal hsa-miR-140-3p could be recognized as a stage specific biomarker. The data also supports the computational prediction that hsa-miR-140-3p is involved in GABA and/or choline associated regulatory network (Ntoumou *et al*, 2017; Zhao *et al*, 2012). Overall, the analysis of exosomal miRNA could become a promising diagnostic strategy to monitor withdrawal symptoms in the context of SUDs.

Furthermore, an increase of exosomal hsa-miR-744-5p has been identified in patients with SUDs at the short-term withdrawal stage and it is positively correlated with serotonin concentration in the serum in both HDPs and MDPs (**Figure S5**). Previous studies have also shown that hsa-miR-744-5p inhibits cell proliferation and invasion in pituitary oncocytoma and epithelial ovarian cancer (Chen *et al*, 2019a; Krokker *et al*, 2019; Zhao *et al*, 2020). Herein, we performed bioinformatical analysis and identified genes PSENEN, FDX1L, VPS37D, SPTBN4, PPP5C, and FAM57B, as potential targets of hsa-miR-744-5p (**Figure 4K**). PSENEN, as a key regulator of the gamma-secretase complex which is involved in the production of the amyloid beta 42 peptides, has been reported to be associated with late onset Alzheimer’s disease (Jia *et al*, 2007; Nam *et al*, 2011). In addition, SPTBN4 disorder is characterized by severe-to-profound developmental delay and/or intellectual disability (Wang *et al*, 2018), while DNA methylation-associated silencing in SPTBN4 was also identified in patients with Alzheimer’s disease (Sanchez-Mut *et al*, 2013). Therefore, we speculate that hsa-744-5p may regulate key neuronal genes PSENEN and SPTBN4 to suppress neurogenesis and contribute to the neurological and/or psychological symptoms observed in patients with SUDs.

More importantly, these above findings point towards unknown mechanisms in patients with SUDs that dysregulated exosomal miRNAs may be functioning directly on CNS. Several studies have shown the capability of exosomes to cross the BBB and communicate between the peripheral circulation and the CNS (Chen *et al*., 2016; Saeedi *et al*., 2019). In the present study, we labeled exosomes from human serum with DiR, injected them via the tail vein of C57/BL6 mice, and imaged them 24 hours after injection using an *in vivo* imaging system. Of note, DiR-labeled exosomes were successfully captured in the brain and other organs (**Figure S6A-C**). We further compared the ability of exosomes crossing the BBB in the mice with or without LPS treatment. Interestingly, a significant increase in DiR-labeled exosomes was captured and measured in the brain from LPS treated mice (**Figure S6D**), suggesting that we are indeed tracking labeled exosomes crossing the BBB and more exosomes crossing the BBB when permeability is disrupted by LPS treatment and inflammation (Banks *et al*, 2015; Banks *et al*, 2020). However, the biogenesis of exosomes across endothelial cells is a relatively unexplored area and it is unknown whether the changes in miRNAs between the CNS and the peripheral circulation could be due to off-loading miRNA cargo for endothelial regulation or other possible factors. Future studies may unravel how exosomes change through membrane barriers of the brain and various organs in the periphery before circulating in the bloodstream.

The fact, although both heroin and methamphetamine are highly addictive and dangerous drugs, they belong to two different categories of addictive substances, opiates and stimulants, respectively, leading to distinct major effects and their acting through different mechanisms. Surprisingly, we observed a large overlap in heroin and methamphetamine induced neurotransmitter and exosomal miRNA changes during withdrawal. Thus, future longitudinal studies with larger sample sizes will be needed to uncover the subtle differences between heroin and methamphetamine induced changes. The presented study also entails limitations due to the protocol and the patient chosen. Due to the protocol applied, we only profiled the exosomal miRNAs and the neurotransmitters present in plasma. However, the origin of these exosomes with dysregulated miRNAs has not been identified. Further biological studies and more laborious techniques such as proteomics should address the origin of these exosomes with dysregulated miRNAs.

In conclusion, to the best of our knowledge, this study has the largest sample size in the field to date and performed in a controlled setting to correlate the severity of substance withdrawal stress with alterations in plasma neurotransmitter and exosomal miRNA expression profiles. By using UPLC-MS/MS and high-throughput sequencing approaches and by including an extensive functional validation analysis, we have identified a set of neurotransmitters associated exosomal miRNAs which may elucidate the molecular mechanism of how the peripheral circulation communicates with the CNS and contributes to the symptoms associated with substance dependence and withdrawal. We also demonstrated plausible mechanistic links in an *in vitro* neuronal model and the signaling pathways in which they operate. Overall, these data add to a growing literature that implicates plasma exosomal miRNAs as functional mediators of neuronal function and development processes of withdrawal symptoms.

## Data Availability

The clinical data of the patients used to support the findings of this study are included within the article. The original sequencing data of the microRNA used to support the findings of this study are available at PRJNA667326 of NCBI Sequence Read Archive (SRA).

## Author contributions

JH.Y. and KH.W. designed the study and supervised the project. Y.X., FR.C., ZY.Z., ZR.X., YR.M., L.Z., YQ.K., J.L., Y.W. and M.Z. collected the data. JH.Y., K.S., HJ.W. and QY.P. did the data analysis and interpretation. JH.Y. took the lead in writing the manuscript. T.T. and T.W.R. discussed the results and contributed to the final manuscript. All authors reviewed the report and approved the final version.

## Acknowledgments

Foremost, we would like to thank Yongjin Zhang and Limei Cao for their expert technique assistance. This work was supported by grants from the National Natural Science Foundation of China (Grant No. 3171101074, 81860100, 31860306, and 81870458), Science and Technology Department of Yunnan Province (Grant No. 2018DH006, 2018NS0086, 202001AV070010), Yunnan Province Clinical Research Center (2019ZF012), and Yunling Scholar (Grant No. YLXL20170002).

