## Supplemental Figure 1-6 for "Dynamics of neurotransmitter and extracellular vesicle-derived microRNA landscapes during heroin and methamphetamine withdrawal"

### Supplementary information

#### Supplementary Figures

**Figure S1**

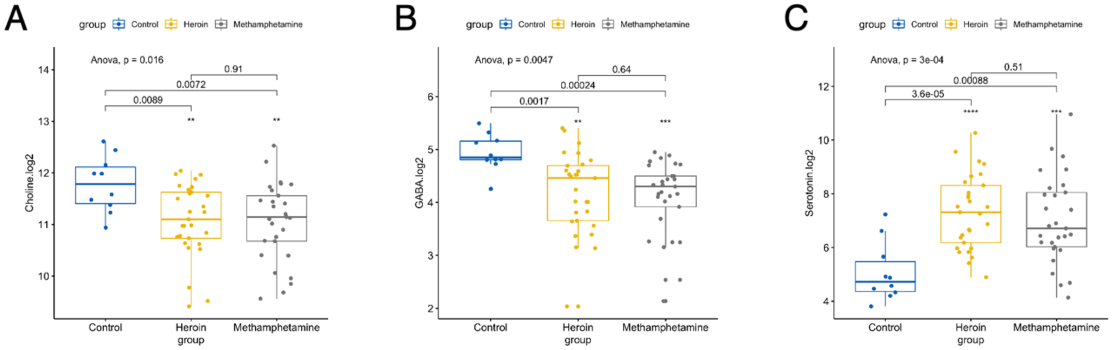


**Figure S1: Differences of serum neurotransmitters in patients with SUDs and healthy controls.** Statistically significances were detected in (A) Choline, (B) GABA, and (C) Serotonin levels between patients with SUDs and healthy controls.

### Figure S2

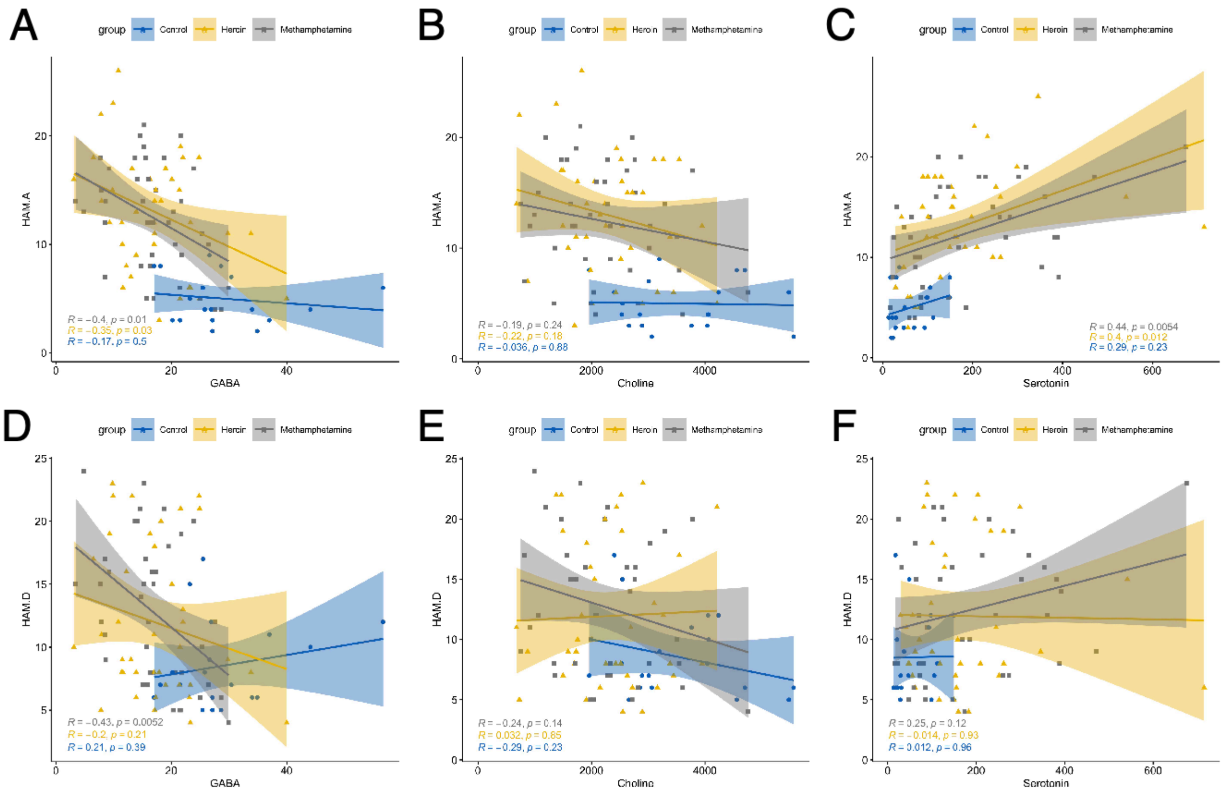


**Figure S2: Sparse canonical correlation analysis.** Sparse canonical correlation analyses were carried out in (A, B, C) HAM-A and (D, E, F) HAM-D scales versus GABA, Choline and Serotonin levels in patients with SUDs.

**Figure S3**

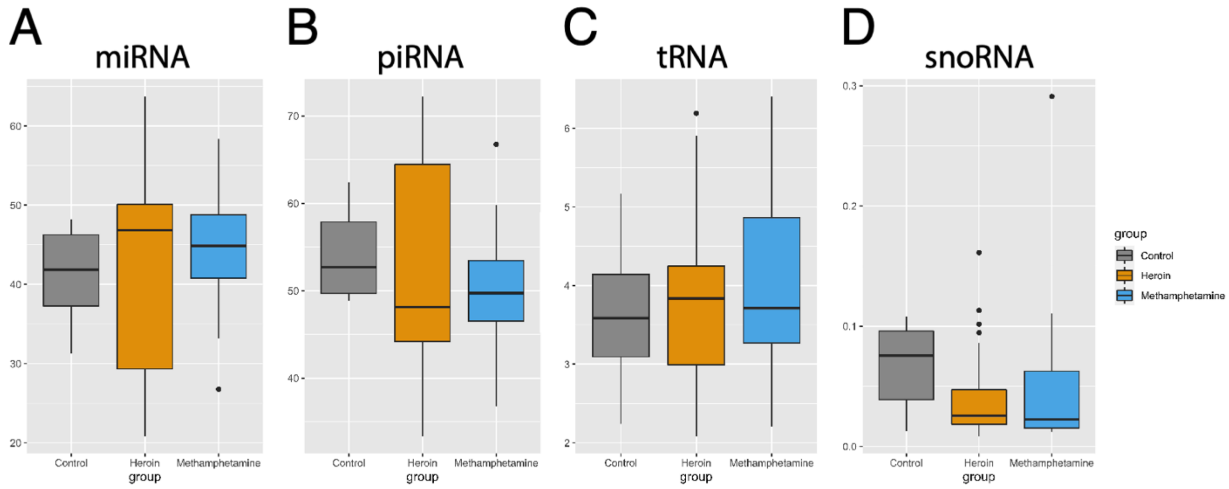


**Figure S3: Box plot representing percentage of reads assigned to different RNA species from sequencing,** including (A) miRNA, (B) piRNA, (C) tRNA and (D) snoRNA.

**Figure S4**

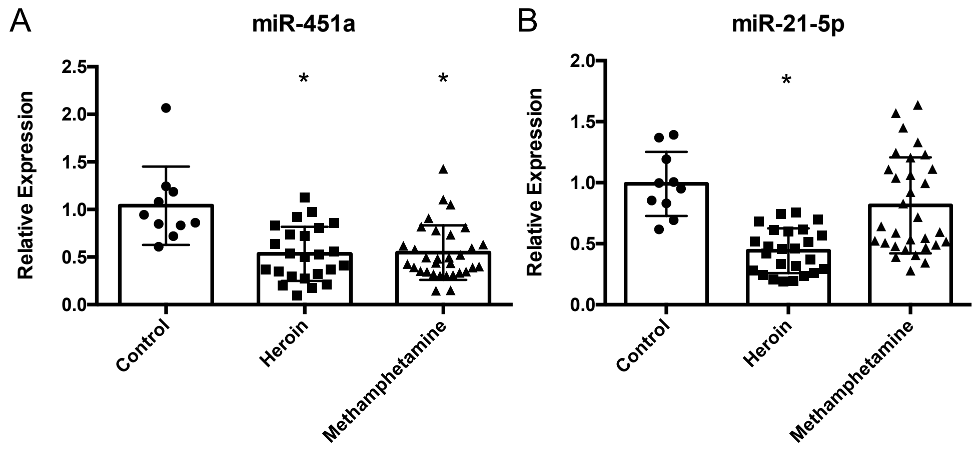


**Figure S4: qPCR validation of identified miRNAs using an independent cohort.** The expressions of (A) miR-451a and (B) miR-21-5p were validated using qPCR. Error bars represent the standard error of the mean of the samples analysed. * represents $p < 0.05$.

**Figure S5**

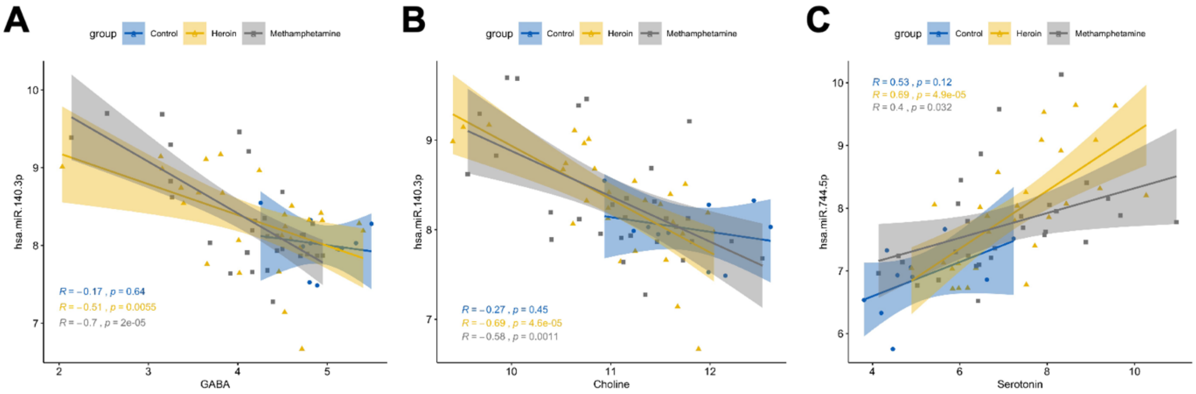


**Figure S5: Sparse canonical correlation analysis.** (A, B) Sparse canonical correlation analyses were carried out in exosomal hsa-miR-140-3p versus GABA (A) and Choline (B), respectively, in patients with SUDs and healthy controls. (C) Sparse canonical correlation analyses were carried out in exosomal hsa-miR-744-5p versus Serotonin in patients with SUDs and healthy controls.

#### Figure S6

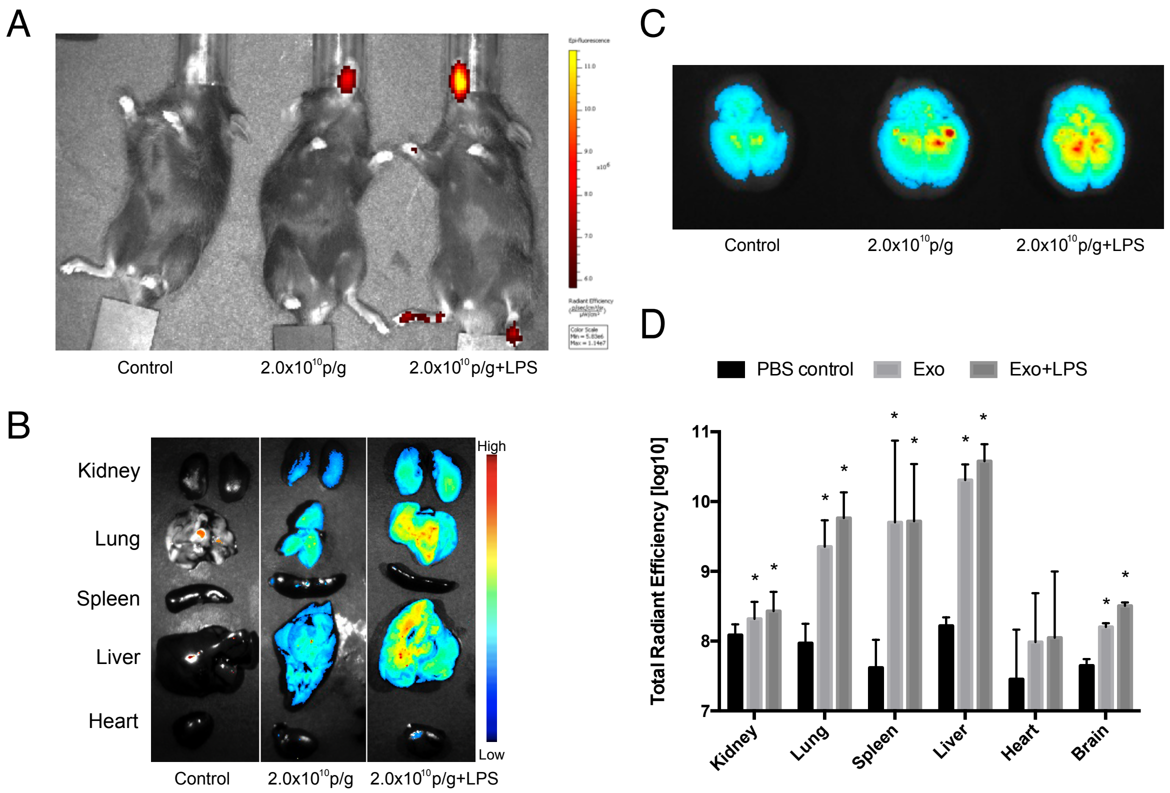


**Figure S6: Distribution of fluorescent lipophilic tracer DiR-labelled exosomes in mice.** (A) Lipopolysaccharide (LPS) or PBS treated C57BL/6 mice (*n* = 3/group) were used and freshly purified DiR-labeled exosomes were injected through the tail vein for intravenous injections. The biodistribution of labeled exosomes was examined using 2.0 x $10^{10}$ particles/gram body weight (p/g); (B,C) DiR-labeled exosomes were successfully captured in the brain and other organs in mice; (D) A significant increase in DiR-labeled exosomes was captured and measured in the brain and other organs from LPS treated mice (n=3).
