## Supplemental Table 1-9 for "Dynamics of neurotransmitter and extracellular vesicle-derived microRNA landscapes during heroin and methamphetamine withdrawal"

### Supplementary Table 1 Characteristics of study participants from the discovery cohort.

| | *mean* | *mean Heroin subgroup* | | | ***p*** | ***fdr.p*** | ***p*** | ***fdr.p*** | ***p*** | ***fdr.p*** | ***p*** | ***fdr.p*** | ***p*** | ***fdr.p*** | ***p*** | ***fdr.p*** |
|---|---|---|---|---|---|---|---|---|---|---|---|---|---|---|---|---|
| | **Control (n=10)** | **T1 (n=10)** | **T2 (n=10)** | **T3 (n=10)** | **Control vs T1** | | **Control vs T2** | | **Control vs T3** | | **T1 vs T2** | | **T2 vs T3** | | **T1 vs T3** | |
| **Age (year)** | 36.71±7.32 | 37.73±7.27 | 37.01±8.21 | 37.00±8.05 | 0.7337 | 1.0000 | 0.8501 | 1.0000 | 0.9097 | 1.0000 | 1.0000 | 1.0000 | 0.9699 | 1.0000 | 0.7337 | 1.0000 |
| **BMI** | 21.36±1.68 | 22.13±1.53 | 21.85±1.81 | 22.18±1.53 | 0.3075 | 0.8178 | 0.8501 | 0.8501 | 0.4274 | 0.8178 | 0.5205 | 0.8178 | 0.5452 | 0.8178 | 0.8501 | 0.8501 |
| **History (year)** | NA | 7.56±2.66 | 7.85±2.42 | 7.99±2.72 | NA | NA | NA | NA | NA | NA | 0.8500 | 0.8501 | 0.8501 | 0.8501 | 0.7913 | 0.8501 |
| **Education*** | 1/6/2/1 | 4/4/1/1 | 3/4/3/0 | 2/4/3/1 | 0.6181 | 1.0000 | 0.5908 | 1.0000 | 0.8363 | 1.0000 | 0.7136 | 1.0000 | 1.0000 | 1.0000 | 0.703 | 1.0000 |
| **Income**** | 0/3/4/3/0 | 2/3/3/1/1 | 2/4/2/1/1 | 1/5/2/2/0 | 0.4984 | 1.0000 | 0.3832 | 1.0000 | 0.6135 | 1.0000 | 1.0000 | 1.0000 | 1.0000 | 1.0000 | 0.7954 | 1.0000 |
| **HAM-A** | 4.9±2.42 | NA | 15.3±5.76 | 9.2±4.32 | NA | NA | **0.0004** | **0.0012** | **0.0166** | **0.0188** | NA | NA | **0.0188** | **0.0188** | NA | NA |
| **HAM-D** | 8.2±3.82 | NA | 14.2±6.51 | 11.7±5.23 | NA | NA | 0.0807 | 0.1211 | **0.0133** | **0.0399** | NA | NA | 0.5435 | 0.5435 | NA | NA |
| | *mean* | *mean Methamphetamine subgroup* | | | ***p*** | ***fdr.p*** | ***p*** | ***fdr.p*** | ***p*** | ***fdr.p*** | ***p*** | ***fdr.p*** | ***p*** | ***fdr.p*** | ***p*** | ***fdr.p*** |
| | **Control (n=10)** | **T1 (n=10)** | **T2 (n=10)** | **T3 (n=10)** | **Control vs T1** | | **Control vs T2** | | **Control vs T3** | | **T1 vs T2** | | **T2 vs T3** | | **T1 vs T3** | |
| **Age (year)** | 36.71±7.32 | 36.07±7.77 | 36.93±7.81 | 36.73±8.38 | 0.9097 | 0.9097 | 0.8501 | 0.9097 | 0.9097 | 0.9097 | 0.7913 | 0.9097 | 0.8501 | 0.9097 | 0.7913 | 0.9097 |
| **BMI** | 21.36±1.68 | 21.56±1.69 | 21.70±1.33 | 21.42±1.52 | 0.8501 | 0.9698 | 0.7623 | 0.9698 | 0.7623 | 0.9698 | 0.9698 | 0.9698 | 0.5708 | 0.9698 | 0.5205 | 0.9698 |
| **History (year)** | NA | 7.4±2.25 | 7.34±3.35 | 7.63±2.93 | NA | NA | NA | NA | NA | NA | 0.8797 | 0.8797 | 0.5708 | 0.8797 | 0.7911 | 0.8797 |
| **Education*** | 1/6/2/1 | 3/3/3/1 | 3/3/4/0 | 3/5/2/0 | 0.6454 | 1.0000 | 0.2862 | 1.0000 | 0.82 | 1.0000 | 1.0000 | 1.0000 | 0.8484 | 1.0000 | 0.8484 | 1.0000 |
| **Income**** | 0/3/4/3/0 | 1/4/2/3/0 | 1/4/2/2/1 | 3/3/1/2/1 | 0.8484 | 1.0000 | 0.7727 | 1.0000 | 0.2525 | 1.0000 | 1.0000 | 1.0000 | 0.9182 | 1.0000 | 0.7954 | 1.0000 |
| **HAM-A** | 4.9±2.42 | NA | 13.6±3.86 | 9±5.31 | NA | NA | **0.0006** | **0.0019** | **0.0391** | **0.0409** | NA | NA | **0.0409** | **0.0409** | NA | NA |
| **HAM-D** | 8.2±3.82 | NA | 15.2±5.33 | 11.7±6.36 | NA | NA | **0.0050** | **0.0150** | 0.2074 | 0.2074 | NA | NA | 0.1969 | 0.2074 | NA | NA |

Data are mean ± SD. Group differences are evaluated with Kruskal's t test for continuous variables and fisher's test for categorical variables. * Education levels: illiteracy/primary school/middle school/college; ** Income levels: monthly 0~1000¥/1000~3000¥/3000~5000¥/5000~10000¥/10000+¥. HAM-A: Hamilton Rating Scale for Anxiety; HAM-D: Hamilton Depression Rating Scale.

### Supplementary Table 2. Characteristics of study participants from the validation cohort.

| | *mean* | *mean Heroin subgroup* | | | *p* | *fdr.p* | *p* | *fdr.p* | *p* | *fdr.p* | *p* | *fdr.p* | *p* | *fdr.p* | *p* | *fdr.p* |
|---|---|---|---|---|---|---|---|---|---|---|---|---|---|---|---|---|
| | **Control (n=10)** | **T1 (n=9)** | **T2 (n=9)** | **T3 (n=10)** | **Control vs T1** | | **Control vs T2** | | **Control vs T3** | | **T1 vs T2** | | **T2 vs T3** | | **T1 vs T3** | |
| **Age (year)** | 36.03±9.05 | 37.22±7.77 | 36.41±9.77 | 36.87±8.02 | 0.8501 | 1.0000 | 0.9097 | 1.0000 | 0.7913 | 1.0000 | 1.0000 | 1.0000 | 0.9698 | 1.0000 | 0.9699 | 1.0000 |
| **BMI** | 22.01±1.6 | 20.95±2.03 | 21.42±1.93 | 21.76±1.8 | 0.1859 | 0.7913 | 0.5205 | 0.7913 | 0.7913 | 0.7913 | 0.5708 | 0.7913 | 0.6776 | 0.7913 | 0.3847 | 0.7913 |
| **History (year)** | NA | 7.19±2.97 | 6.17±3.19 | 7.01±2.97 | NA | NA | NA | NA | NA | NA | 0.5205 | 0.7808 | 0.4727 | 0.7808 | 0.8501 | 0.8501 |
| **Education*** | 1/4/4/1 | 2/4/3/1 | 2/2/4/2 | 2/4/2/2 | 1.0000 | 1.0000 | 0.7545 | 1.0000 | 0.7545 | 1.0000 | 0.8636 | 1.0000 | 0.9221 | 1.0000 | 1.0000 | 1.0000 |
| **Income**** | 0/4/4/2/0 | 2/4/2/2/0 | 2/3/3/1/1 | 1/5/3/1/0 | 0.5393 | 1.0000 | 0.6658 | 1.0000 | 1.0000 | 1.0000 | 1.0000 | 1.0000 | 0.9091 | 1.0000 | 1.0000 | 1.0000 |
| **HAM-A** | 5.4±2.17 | NA | 15.7±4.11 | 13±2.71 | NA | NA | **0.0003** | **0.0004** | **0.0002** | **0.0004** | NA | NA | 0.0808 | 0.0808 | NA | NA |
| **HAM-D** | 8.8±3.05 | NA | 15.4±6.35 | 10.9±6.62 | NA | NA | **0.0306** | 0.0917 | 0.8791 | 0.8791 | NA | NA | 0.1725 | 0.2587 | NA | NA |
| | *mean* | *mean Methamphetamine subgroup* | | | *p* | *fdr.p* | *p* | *fdr.p* | *p* | *fdr.p* | *p* | *fdr.p* | *p* | *fdr.p* | *p* | *fdr.p* |
| | **Control (n=10)** | **T1 (n=9)** | **T2 (n=9)** | **T3 (n=10)** | **Control vs T1** | | **Control vs T2** | | **Control vs T3** | | **T1 vs T2** | | **T2 vs T3** | | **T1 vs T3** | |
| **Age (year)** | 36.03±9.05 | 36.36±8.36 | 35.95±7.9 | 36.54±9.78 | 1.0000 | 1.0000 | 0.9699 | 1.0000 | 1.0000 | 1.0000 | 0.9097 | 1.0000 | 0.7337 | 1.0000 | 0.7913 | 1.0000 |
| **BMI** | 22.01±1.6 | 21.85±1.74 | 21.86±1.01 | 21.02±0.84 | 0.9698 | 0.9698 | 0.9097 | 0.9698 | 0.1986 | 0.3972 | 0.7911 | 0.9698 | 0.0639 | 0.2270 | 0.0757 | 0.2270 |
| **History (year)** | NA | 7.29±2.99 | 7.62±2.12 | 6.26±2.39 | NA | NA | NA | NA | NA | NA | 0.6500 | 0.7337 | 0.2413 | 0.7239 | 0.7337 | 0.7337 |
| **Education*** | 1/4/4/1 | 3/2/3/2 | 2/4/2/2 | 2/3/4/1 | 0.6704 | 1.0000 | 0.7545 | 1.0000 | 1.0000 | 1.0000 | 0.8701 | 1.0000 | 0.8636 | 1.0000 | 1.0000 | 1.0000 |
| **Income**** | 0/4/4/2/0 | 1/5/2/2/0 | 2/3/0/3/2 | 2/3/3/2/0 | 0.8363 | 0.8636 | 0.08236 | 0.4942 | 0.7348 | 0.8636 | 0.4484 | 0.8636 | 0.3647 | 0.8636 | 0.8636 | 0.8636 |
| **HAM-A** | 5.4±2.17 | NA | 16.1±3.87 | 11±3.86 | NA | NA | **0.0002** | **0.0005** | **0.0039** | **0.0058** | NA | NA | **0.0166** | **0.0166** | NA | NA |
| **HAM-D** | 8.8±3.05 | NA | 16.3±4.37 | 11.3±5.93 | NA | NA | **0.0014** | **0.0043** | 0.5681 | 0.5681 | NA | NA | 0.0634 | 0.0951 | NA | NA |

Data are mean ± SD. Group differences are evaluated with Kruskal's t test for continuous variables and fisher's test for categorical variables. * Education levels: illiteracy/primary school/middle school/college; ** Income levels: monthly 0~1000¥/1000~3000¥/3000~5000¥/5000~10000¥/10000+¥. HAM-A: Hamilton Rating Scale for Anxiety; HAM-D: Hamilton Depression Rating Scale.

**Supplementary Table 3** Detailed information of the 14 neurotransmitters in UPLC-MS/MS

| No. | Metabolites | HMDB ID |
|---|---|---|
| 1 | γ-Aminobutyric acid | HMDB0000112 |
| 2 | Glutamine | HMDB0003423 |
| 3 | Glutamate | HMDB0000148 |
| 4 | Glutathione | HMDB0000125 |
| 5 | Tryptophan | HMDB0000929 |
| 6 | Choline | HMDB0000097 |
| 7 | Phosphocholine | HMDB0001565 |
| 8 | Tyramine | HMDB0000306 |
| 9 | Serotonin | HMDB0000259 |
| 10 | Myo-inositol | HMDB0000211 |
| 11 | Dopamine | HMDB0000073 |
| 12 | 5-Hydroxyindole acetic acid | HMDB0001855 |
| 13 | Epinephrine | HMDB0000068 |
| 14 | Kynurenine | HMDB0000684 |

**Supplementary Table 4. Detailed information of the neurotransmitters isolated from peripheral blood plasma of HC, HDPs and MDPs from the validation cohort**

| Metabolites | *mean* | *mean* | *mean* | *p* | *fdr.p* | *p* | *fdr.p* | *p* | *fdr.p* |
|---|---|---|---|---|---|---|---|---|---|
| | **Control (n=10)** | **Heroin (n=30)** | **Methamphetamine (n=30)** | **Control vs Heroin** | | **Control vs Methamphetamine** | | **Heroin vs Methamphetamine** | |
| **Choline** | 3639.23±978.58 | 2484.73±604.82 | 2341.57±641.27 | **0.0008*** | **0.0012** | **0.0005*** | **0.0012** | 0.2739 | 0.2739 |
| **GABA** | 28.38±11.39 | 17.70±6.32 | 17.24±4.17 | **0.0006*** | **0.0009** | **0.0002*** | **0.0006** | 0.9117 | 0.9117 |
| **Glutamic acid** | 47698.77±35785.73 | 37640.50±18612.48 | 30323.27±11312.50 | 1.0000 | 1.0000 | 0.1949 | 0.2924 | **0.0232*** | 0.0696 |
| **Glutamine** | 10040.83±6513.06 | 7406.37±1668.19 | 8005.35±1850.10 | 0.1221 | 0.3663 | 0.7194 | 0.7194 | 0.2838 | 0.4257 |
| **Kynurenine** | 1759.80±723.00 | 1363.88±646.31 | 1380.89±645.64 | 0.1147 | 0.1720 | 0.1147 | 0.1720 | 0.7731 | 0.7731 |
| **Serotonin** | 79.08±38.78 | 317.00±298.67 | 317.30±280.52 | **0.0001*** | **0.0003** | **0.0003*** | **0.0004** | 0.6843 | 0.6843 |
| **Tryptophan** | 24921.68±4269.84 | 20620.83±4911.53 | 23067.06±5320.18 | **0.0255*** | 0.0765 | 0.2675 | 0.2675 | 0.1154 | 0.1731 |

Data are mean ± SD. Group differences are evaluated with Kruskal's t test for continuous variables and fisher's test for categorical variables.

### Supplementary Table 5. Subgroup analysis of 7 neurotransmitters isolated from peripheral blood plasma of HC, HDPs and MDPs.

| Metabolites | *mean* | *mean Heroin subgroup* | | | *p* | *fdr.p* | *p* | *fdr.p* | *p* | *fdr.p* | *p* | *fdr.p* | *p* | *fdr.p* | *p* | *fdr.p* |
|---|---|---|---|---|---|---|---|---|---|---|---|---|---|---|---|---|
| | **Control (n=10)** | **T1 (n=10)** | **T2 (n=10)** | **T3 (n=10)** | **Control vs T1** | | **Control vs T2** | | **Control vs T3** | | **T1 vs T2** | | **T2 vs T3** | | **T1 vs T3** | |
| **Glutamine** | 10040.83± 6513.06 | 6550.29±1433.48 | 7491.73±1594.10 | 8177.11±1699.56 | **0.0113** | 0.0678 | 0.2413 | 0.3620 | 1.0000 | 1.0000 | 0.1212 | 0.2424 | 0.3847 | 0.4616 | **0.0452** | 0.1355 |
| **Tryptophan** | 24921.68± 4269.84 | 18988.44± 6345.78 | 20849.91± 4313.04 | 22024.15± 3694.80 | 0.0539 | 0.1617 | **0.0452** | 0.1617 | 0.1620 | 0.3239 | 0.6232 | 0.6232 | 0.5708 | 0.6232 | 0.2730 | 0.4096 |
| **Choline** | 3639.23±978.58 | 2615.30±609.90 | 2274.89±638.28 | 2564.01±569.62 | **0.0211** | **0.0422** | **0.0022** | **0.0132** | **0.0073** | **0.0219** | 0.1211 | 0.1816 | 0.2730 | 0.3276 | 0.5204 | 0.5204 |
| **GABA** | 28.38±11.39 | 20.33±8.28 | 14.07±5.03 | 18.71±3.35 | **0.0211** | **0.0423** | **0.0008** | **0.0046** | **0.0091** | **0.0273** | 0.0890 | 0.1068 | 0.0539 | 0.0809 | 0.7337 | 0.7337 |
| **Glutamic acid** | 47698.77± 35785.73 | 43557.35± 30121.80 | 31334.81 ±7359.47 | 38029.35± 8457.26 | 0.9698 | 0.9698 | 0.4727 | 0.7808 | 0.5205 | 0.7808 | 0.3447 | 0.7808 | 0.1041 | 0.6247 | 0.7337 | 0.8805 |
| **Serotonin** | 79.08±38.78 | 531.14±396.70 | 278.38±200.17 | 141.48±56.23 | **0.0002** | **0.0015** | **0.0013** | **0.0039** | **0.0173** | **0.0259** | 0.1041 | 0.1041 | 0.0539 | 0.0647 | **0.0022** | **0.0044** |
| **Kynurenine** | 1759.80±723.00 | 1273.71±527.54 | 1510.07±876.60 | 1307.87±512.81 | 0.1405 | 0.4214 | 0.4727 | 0.6246 | 0.1212 | 0.4214 | 0.5205 | 0.6246 | 0.4727 | 0.6246 | 1.0000 | 1.0000 |

| Metabolites | *mean* | *mean Methamphetamine subgroup* | | | *p* | *fdr.p* | *p* | *fdr.p* | *p* | *fdr.p* | *p* | *fdr.p* | *p* | *fdr.p* | *p* | *fdr.p* |
|---|---|---|---|---|---|---|---|---|---|---|---|---|---|---|---|---|
| | **Control (n=10)** | **T1 (n=10)** | **T2 (n=10)** | **T3 (n=10)** | **Control vs T1** | | **Control vs T2** | | **Control vs T3** | | **T1 vs T2** | | **T2 vs T3** | | **T1 vs T3** | |
| **Glutamine** | 10040.83± 6513.06 | 7041.57±1466.38 | 7862.24±1363.41 | 9112.23±2145.69 | 0.0757 | 0.2107 | 0.9698 | 0.9698 | 0.4274 | 0.5128 | 0.1212 | 0.2107 | 0.1405 | 0.2107 | 0.0312 | 0.1873 |
| **Tryptophan** | 24921.68± 4269.84 | 21431.59± 4630.92 | 23593.16± 6200.37 | 24176.45± 5163.90 | 0.0757 | 0.4540 | 0.7337 | 0.8805 | 0.6232 | 0.8805 | 0.3075 | 0.8805 | 0.9097 | 0.9097 | 0.4727 | 0.8805 |
| **Choline** | 3639.23±978.58 | 2572.38±591.69 | 1877.25±492.63 | 2575.08±610.70 | **0.0140** | **0.0207** | **0.0006** | **0.0035** | **0.0140** | **0.0207** | **0.0173** | **0.0207** | **0.0113** | **0.0207** | 0.9097 | 0.9097 |
| **GABA** | 28.38±11.39 | 18.55±4.33 | 14.18±3.21 | 19.00±3.36 | **0.0140** | **0.0280** | **0.0002** | **0.0011** | **0.0091** | **0.0273** | **0.0376** | **0.0452** | **0.0257** | **0.0386** | 0.5708 | 0.5708 |
| **Glutamic acid** | 47698.77± 35785.73 | 29036.45± 7561.09 | 30838.97± 12857.25 | 31094.38± 13677.77 | 0.3075 | 0.6894 | 0.3447 | 0.6894 | 0.2730 | 0.6894 | 0.7913 | 1.0000 | 0.9698 | 1.0000 | 1.0000 | 1.0000 |
| **Serotonin** | 79.08±38.78 | 449.57±420.04 | 317.44±158.63 | 184.91±114.23 | **0.0046** | **0.0138** | **0.0003** | **0.0020** | **0.0257** | **0.0515** | 0.8501 | 0.8501 | 0.0757 | 0.1135 | 0.1212 | 0.1455 |
| **Kynurenine** | 1759.80±723.00 | 1229.36±326.57 | 1210.30±457.73 | 1703.00±924.05 | **0.0452** | 0.2082 | 0.1041 | 0.2082 | 0.9097 | 0.9698 | 0.9698 | 0.9698 | 0.2123 | 0.3184 | 0.0890 | 0.2082 |

Data are mean ± SD. Group differences are evaluated with Kruskal's t test for continuous variables and fisher's test for categorical variables.

**Supplementary Table 6. Sequencing Quality Statist**

| Sample | Number of reads | | | | | Number of miRNA | |
|---|---|---|---|---|---|---|---|
| | Raw reads | Length trimmed | Quality filtered | N trimmed | Clean reads | Known | Putative novel |
| All samples | 1575073176 | 625036325 | 625036325 | 625030902 | 625030902 | 1523 | 157 |
| C.01 | 28951357 | 6993344 | 6993344 | 6993340 | 6993340 | 612 | 22 |
| C.02 | 24323530 | 5710597 | 5710597 | 5710596 | 5710596 | 552 | 21 |
| C.03 | 25745696 | 8515690 | 8515690 | 8515689 | 8515689 | 598 | 24 |
| C.04 | 23373769 | 8358123 | 8358123 | 8358122 | 8358122 | 662 | 26 |
| C.05 | 20854033 | 5018514 | 5018514 | 5018512 | 5018512 | 436 | 24 |
| C.06 | 23011904 | 8686982 | 8686982 | 8686979 | 8686979 | 646 | 27 |
| C.07 | 19440842 | 4939177 | 4939177 | 4939170 | 4939170 | 468 | 16 |
| C.08 | 23087794 | 8468396 | 8468396 | 8468392 | 8468392 | 592 | 31 |
| C.09 | 25512976 | 7368351 | 7368351 | 7368347 | 7368347 | 618 | 21 |
| C.10 | 21970702 | 7339262 | 7339262 | 7339252 | 7339252 | 614 | 24 |
| H1.1 | 22961536 | 9956030 | 9956030 | 9956030 | 9956030 | 698 | 35 |
| H1.2 | 19931639 | 8877600 | 8877600 | 8877599 | 8877599 | 613 | 21 |
| H1.3 | 20466688 | 8784872 | 8784872 | 8784868 | 8784868 | 396 | 12 |
| H1.4 | 23550416 | 9231064 | 9231064 | 9231064 | 9231064 | 492 | 16 |
| H1.5 | 22312916 | 9901625 | 9901625 | 9901625 | 9901625 | 580 | 23 |
| H1.6 | 21742872 | 9960966 | 9960966 | 9960966 | 9960966 | 609 | 29 |
| H1.7 | 23151988 | 12515142 | 12515142 | 12515142 | 12515142 | 653 | 23 |
| H1.8 | 24137578 | 7783240 | 7783240 | 7783240 | 7783240 | 482 | 20 |
| H1.9 | 20629386 | 8998094 | 8998094 | 8998094 | 8998094 | 619 | 29 |
| H2.1 | 18770486 | 6947738 | 6947738 | 6947738 | 6947738 | 438 | 12 |
| H2.2 | 19452501 | 8092496 | 8092496 | 8090758 | 8090758 | 360 | 10 |
| H2.3 | 30102744 | 13926937 | 13926937 | 13926937 | 13926937 | 510 | 21 |
| H2.4 | 20519446 | 9806286 | 9806286 | 9806286 | 9806286 | 528 | 19 |
| H2.5 | 20054726 | 8762403 | 8762403 | 8762403 | 8762403 | 655 | 21 |
| H2.6 | 20632304 | 9825993 | 9825993 | 9825990 | 9825990 | 597 | 23 |
| H2.7 | 20027723 | 10135235 | 10135235 | 10135235 | 10135235 | 479 | 15 |
| H2.8 | 17216335 | 6483377 | 6483377 | 6483376 | 6483376 | 489 | 23 |
| H2.9 | 20277871 | 7682860 | 7682860 | 7682860 | 7682860 | 501 | 18 |
| H2.10 | 18488431 | 7421336 | 7421336 | 7421334 | 7421334 | 486 | 14 |
| H3.1 | 20163084 | 8897080 | 8897080 | 8897060 | 8897060 | 321 | 9 |
| H3.2 | 33321634 | 10892910 | 10892910 | 10892910 | 10892910 | 452 | 14 |
| H3.3 | 23960767 | 12740051 | 12740051 | 12740051 | 12740051 | 523 | 25 |
| H3.4 | 22811526 | 10904450 | 10904450 | 10904450 | 10904450 | 438 | 20 |

| | | | | | | | |
|---|---|---|---|---|---|---|---|
| H3.5 | 21481696 | 10888465 | 10888465 | 10888465 | 10888465 | 540 | 17 |
| H3.6 | 25475566 | 11963806 | 11963806 | 11963806 | 11963806 | 484 | 16 |
| H3.7 | 22163682 | 10646737 | 10646737 | 10646737 | 10646737 | 484 | 11 |
| H3.8 | 19209212 | 9263671 | 9263671 | 9263671 | 9263671 | 435 | 13 |
| H3.9 | 20427766 | 8026759 | 8026759 | 8026753 | 8026753 | 384 | 13 |
| H3.10 | 23919430 | 10474858 | 10474858 | 10474858 | 10474858 | 357 | 9 |
| M1.1 | 20872402 | 7228699 | 7228699 | 7228699 | 7228699 | 452 | 17 |
| M1.2 | 20097637 | 7595432 | 7595432 | 7595432 | 7595432 | 455 | 21 |
| M1.3 | 18206676 | 6112801 | 6112801 | 6112801 | 6112801 | 577 | 24 |
| M1.4 | 21754137 | 6015848 | 6015848 | 6015848 | 6015848 | 462 | 13 |
| M1.5 | 26026823 | 8548506 | 8548506 | 8548506 | 8548506 | 588 | 25 |
| M1.6 | 23213882 | 9173126 | 9173126 | 9173126 | 9173126 | 696 | 25 |
| M1.7 | 19352539 | 6250781 | 6250781 | 6250781 | 6250781 | 553 | 27 |
| M1.8 | 24561326 | 7055055 | 7055055 | 7055055 | 7055055 | 649 | 25 |
| M1.9 | 26054761 | 8762625 | 8762625 | 8762625 | 8762625 | 675 | 33 |
| M1.10 | 31357518 | 9885420 | 9885420 | 9885420 | 9885420 | 577 | 27 |
| M2.1 | 18686291 | 7814755 | 7814755 | 7814753 | 7814753 | 516 | 20 |
| M2.2 | 19003637 | 4690820 | 4690820 | 4690194 | 4690194 | 380 | 10 |
| M2.3 | 19159441 | 8429956 | 8429956 | 8429955 | 8429955 | 494 | 18 |
| M2.4 | 19387191 | 7719314 | 7719314 | 7719314 | 7719314 | 432 | 9 |
| M2.5 | 17760736 | 6064302 | 6064302 | 6064302 | 6064302 | 442 | 13 |
| M2.6 | 18348826 | 8840170 | 8840170 | 8839600 | 8839600 | 560 | 21 |
| M2.7 | 19776356 | 9027219 | 9027219 | 9026441 | 9026441 | 591 | 36 |
| M2.8 | 19283791 | 8803941 | 8803941 | 8803140 | 8803140 | 590 | 27 |
| M2.9 | 19239913 | 10264909 | 10264909 | 10264909 | 10264909 | 664 | 33 |
| M2.10 | 18085579 | 8621063 | 8621063 | 8620323 | 8620323 | 438 | 26 |
| M3.1 | 25300934 | 11235627 | 11235627 | 11235601 | 11235601 | 380 | 16 |
| M3.2 | 28112666 | 12467506 | 12467506 | 12467485 | 12467485 | 410 | 11 |
| M3.3 | 20079964 | 9616208 | 9616208 | 9616208 | 9616208 | 559 | 20 |
| M3.4 | 25344093 | 8302942 | 8302942 | 8302935 | 8302935 | 480 | 17 |
| M3.5 | 38967719 | 13921590 | 13921590 | 13921590 | 13921590 | 474 | 19 |
| M3.6 | 16743902 | 7661639 | 7661639 | 7661630 | 7661630 | 397 | 16 |
| M3.7 | 24695439 | 9676900 | 9676900 | 9676885 | 9676885 | 546 | 18 |
| M3.8 | 25928401 | 14050022 | 14050022 | 14050022 | 14050022 | 516 | 18 |
| M3.9 | 23066766 | 8917392 | 8917392 | 8917384 | 8917384 | 505 | 18 |
| M3.10 | 31323165 | 12397377 | 12397377 | 12397377 | 12397377 | 463 | 22 |

Supplementary Table 7 Length distribution analysis of the miRNAs in all samples

| ID | group | 18 | 19 | 20 | 21 | 22 | 23 | 24 | 25 | 26 | 27 | 28 | 29 | 30 | 31 | 32 | total reads |
|---|---|---|---|---|---|---|---|---|---|---|---|---|---|---|---|---|---|
| C.01 | Control | 650043 | 532467 | 628105 | 888367 | 1770639 | 613854 | 385832 | 273844 | 227456 | 228964 | 183021 | 165546 | 164801 | 135373 | 145028 | 6993340 |
| C.02 | Control | 534072 | 439579 | 502974 | 768574 | 1408844 | 489498 | 314626 | 236057 | 193133 | 183385 | 151276 | 138466 | 135793 | 106426 | 107893 | 5710596 |
| C.03 | Control | 646552 | 527933 | 772748 | 1340507 | 2653662 | 677943 | 387039 | 286215 | 230750 | 213093 | 179690 | 164440 | 161290 | 134569 | 139258 | 8515689 |
| C.04 | Control | 477425 | 437886 | 959468 | 1249250 | 2907923 | 700784 | 318813 | 235732 | 198545 | 179231 | 155995 | 148536 | 148548 | 121751 | 118235 | 8358122 |
| C.05 | Control | 446295 | 402612 | 453875 | 604271 | 1033860 | 450850 | 308066 | 239827 | 192791 | 182461 | 156708 | 146780 | 151495 | 121327 | 127294 | 5018512 |
| C.06 | Control | 646289 | 513768 | 711379 | 1177638 | 3036447 | 739642 | 408957 | 276031 | 220075 | 202023 | 168074 | 157781 | 160382 | 128278 | 140215 | 8686979 |
| C.07 | Control | 423937 | 376288 | 441266 | 628269 | 1246088 | 433319 | 284033 | 208173 | 165561 | 158203 | 132844 | 121533 | 121970 | 96219 | 101467 | 4939170 |
| C.08 | Control | 561027 | 504944 | 713230 | 1228081 | 2769359 | 721781 | 415608 | 287105 | 234394 | 228915 | 182299 | 169066 | 170485 | 133744 | 148354 | 8468392 |
| C.09 | Control | 479728 | 421269 | 620857 | 1012013 | 2539965 | 629098 | 356677 | 241263 | 192578 | 179110 | 151668 | 145882 | 151190 | 124536 | 122513 | 7368347 |
| C.10 | Control | 525877 | 466738 | 646425 | 955406 | 2085276 | 569991 | 375666 | 286932 | 250162 | 240177 | 212625 | 203700 | 194502 | 164562 | 161213 | 7339252 |
| H1.1 | Heroin | 1660736 | 548416 | 731861 | 1126337 | 2660349 | 818166 | 483192 | 323657 | 280477 | 328455 | 239075 | 211319 | 201063 | 166169 | 176758 | 9956030 |
| H1.2 | Heroin | 1580145 | 556901 | 612777 | 916087 | 1773159 | 725396 | 491333 | 361411 | 326885 | 347016 | 278321 | 252133 | 244204 | 207024 | 204807 | 8877599 |
| H1.3 | Heroin | 1192259 | 609663 | 676979 | 914863 | 1262139 | 695119 | 549158 | 483613 | 421267 | 416559 | 358109 | 338278 | 332592 | 275071 | 259199 | 8784868 |
| H1.4 | Heroin | 1676745 | 644714 | 658591 | 872768 | 1418045 | 700639 | 533535 | 431686 | 389158 | 392804 | 335224 | 320375 | 320208 | 271766 | 264806 | 9231064 |
| H1.5 | Heroin | 1778763 | 655053 | 715652 | 1087205 | 2113406 | 752124 | 507031 | 398840 | 343331 | 331640 | 281148 | 260359 | 254001 | 212698 | 210374 | 9901625 |
| H1.6 | Heroin | 2625168 | 597783 | 605433 | 968898 | 1879401 | 735018 | 481268 | 310881 | 286460 | 340612 | 251492 | 226780 | 251312 | 205131 | 195329 | 9960966 |
| H1.7 | Heroin | 3730302 | 631339 | 627165 | 1185372 | 2320921 | 1295824 | 527086 | 346773 | 318219 | 411897 | 276659 | 239797 | 228627 | 190891 | 184270 | 12515142 |
| H1.8 | Heroin | 820471 | 624097 | 650866 | 813826 | 1307189 | 661667 | 498974 | 407892 | 357672 | 354728 | 301171 | 275883 | 260986 | 224735 | 223083 | 7783240 |
| H1.9 | Heroin | 709896 | 518366 | 685562 | 1152876 | 2435536 | 828297 | 516997 | 348868 | 306942 | 352848 | 272019 | 236062 | 230406 | 195914 | 207505 | 8998094 |
| H2.1 | Heroin | 466070 | 426996 | 593748 | 960473 | 1654240 | 599641 | 395123 | 291107 | 249500 | 252052 | 217687 | 220838 | 249014 | 179869 | 191380 | 6947738 |
| H2.2 | Heroin | 532811 | 463937 | 630538 | 994402 | 1527403 | 588205 | 395325 | 299959 | 243759 | 231379 | 202762 | 209223 | 297134 | 1317573 | 156348 | 8090758 |
| H2.3 | Heroin | 777958 | 708606 | 1080424 | 2243336 | 4214314 | 1627559 | 703852 | 435461 | 352795 | 396493 | 303018 | 295732 | 338359 | 205201 | 243829 | 13926937 |
| H2.4 | Heroin | 512511 | 466351 | 780738 | 1559108 | 3201593 | 945944 | 537280 | 291644 | 237340 | 279487 | 202202 | 214505 | 268861 | 136180 | 172542 | 9806286 |
| H2.5 | Heroin | 476399 | 429553 | 673357 | 1372016 | 3001626 | 848810 | 445851 | 248589 | 216837 | 269709 | 178912 | 171225 | 191179 | 109499 | 128841 | 8762403 |
| H2.6 | Heroin | 436899 | 394458 | 835549 | 1459433 | 3616558 | 965405 | 441228 | 254615 | 225723 | 367706 | 218135 | 176505 | 176654 | 119111 | 138011 | 9825990 |
| H2.7 | Heroin | 490589 | 458674 | 698704 | 1778760 | 3015080 | 1851966 | 465742 | 269989 | 200359 | 189014 | 156580 | 157273 | 169002 | 111433 | 122070 | 10135235 |
| H2.8 | Heroin | 326157 | 306903 | 660744 | 1157637 | 2140788 | 552279 | 281771 | 175318 | 140497 | 158363 | 117290 | 110458 | 133160 | 87552 | 134459 | 6483376 |
| H2.9 | Heroin | 471642 | 420126 | 630859 | 1197420 | 2496269 | 728815 | 381668 | 240296 | 198725 | 198079 | 160095 | 149139 | 169388 | 113230 | 127109 | 7682860 |
| H2.10 | Heroin | 489738 | 423127 | 595421 | 1133591 | 2185368 | 796776 | 366630 | 238830 | 194536 | 200630 | 165388 | 164341 | 207059 | 119900 | 139999 | 7421334 |
| H3.1 | Heroin | 617766 | 570861 | 898572 | 1395345 | 2522400 | 703807 | 432088 | 320233 | 270372 | 260420 | 212880 | 195137 | 182745 | 151579 | 162855 | 8897060 |
| H3.2 | Heroin | 902518 | 795183 | 1014848 | 1615809 | 2905310 | 929984 | 578838 | 401789 | 330787 | 320571 | 256331 | 232856 | 227886 | 186748 | 193452 | 10892910 |
| H3.3 | Heroin | 628665 | 577118 | 1075460 | 2269143 | 4614665 | 1125427 | 558060 | 338802 | 274296 | 301472 | 231583 | 203791 | 195846 | 164724 | 180999 | 12740051 |
| H3.4 | Heroin | 665600 | 603122 | 1009782 | 1678638 | 3641404 | 864811 | 504279 | 337210 | 288993 | 292482 | 229796 | 212682 | 216552 | 184654 | 174445 | 10904450 |
| H3.5 | Heroin | 534800 | 529306 | 919374 | 3125180 | 3215100 | 954331 | 366489 | 225967 | 190005 | 193933 | 149450 | 132928 | 130166 | 106101 | 115335 | 10888465 |
| H3.6 | Heroin | 659346 | 600298 | 1124045 | 2239406 | 4184974 | 977368 | 482149 | 321838 | 257411 | 264290 | 207279 | 183445 | 177162 | 139190 | 145605 | 11963806 |
| H3.7 | Heroin | 615225 | 557772 | 1009387 | 1810414 | 3584268 | 911998 | 458500 | 306860 | 249550 | 254404 | 206186 | 186845 | 178339 | 147094 | 169895 | 10646737 |
| H3.8 | Heroin | 544098 | 502914 | 813549 | 2053428 | 2729138 | 825121 | 397704 | 259313 | 216297 | 215290 | 168003 | 150542 | 140917 | 115448 | 131909 | 9263671 |
| H3.9 | Heroin | 571230 | 522613 | 811966 | 1394915 | 2132881 | 625873 | 379780 | 281516 | 242046 | 222764 | 193168 | 180015 | 171443 | 144797 | 151746 | 8026753 |
| H3.10 | Heroin | 706629 | 655819 | 1047644 | 1835588 | 2945586 | 928155 | 486022 | 347969 | 287667 | 279848 | 217132 | 202192 | 197585 | 152903 | 184119 | 10474858 |
| M1.1 | Methampl | 705025 | 533021 | 592035 | 807344 | 1295357 | 626808 | 448431 | 366335 | 319743 | 301299 | 267602 | 255968 | 253449 | 226156 | 230126 | 7228699 |
| M1.2 | Methampl | 658207 | 513786 | 616684 | 937343 | 1702280 | 661275 | 448719 | 348165 | 299205 | 289983 | 249657 | 237430 | 235071 | 197354 | 200273 | 7595432 |
| M1.3 | Methampl | 403040 | 371292 | 466689 | 860942 | 1670219 | 571759 | 328126 | 227853 | 193685 | 222120 | 167870 | 151047 | 163008 | 144704 | 170447 | 6112801 |
| M1.4 | Methampl | 598865 | 474943 | 546745 | 736021 | 1342447 | 497685 | 359860 | 265928 | 227144 | 220789 | 174990 | 161519 | 153948 | 126050 | 128914 | 6015848 |
| M1.5 | Methampl | 610050 | 525259 | 709279 | 1170117 | 2526038 | 744049 | 453427 | 315362 | 266398 | 315925 | 223971 | 193019 | 186672 | 150602 | 158338 | 8548506 |
| M1.6 | Methampl | 578510 | 518616 | 777448 | 1433936 | 3003306 | 795974 | 445633 | 295108 | 249303 | 266218 | 195927 | 171959 | 166188 | 128905 | 146095 | 9173126 |
| M1.7 | Methampl | 518132 | 452545 | 538637 | 782642 | 1486471 | 561343 | 367365 | 278522 | 230580 | 227289 | 189314 | 169853 | 168653 | 139379 | 140056 | 6250781 |
| M1.8 | Methampl | 643817 | 531880 | 602770 | 964307 | 1830907 | 649783 | 387000 | 266671 | 217357 | 229589 | 175429 | 158176 | 156836 | 118968 | 121565 | 7055055 |
| M1.9 | Methampl | 643759 | 531823 | 711298 | 1277322 | 2701750 | 792944 | 457721 | 292227 | 237780 | 263480 | 192397 | 171867 | 174843 | 145195 | 168219 | 8762625 |
| M1.10 | Methampl | 764319 | 622701 | 781802 | 1266942 | 2841271 | 868623 | 524771 | 369424 | 325619 | 334346 | 259189 | 241559 | 237036 | 231359 | 216459 | 9885420 |
| M2.1 | Methampl | 460613 | 420391 | 657737 | 1190048 | 2467006 | 739254 | 403362 | 247437 | 207275 | 220684 | 179892 | 168389 | 173203 | 132777 | 146685 | 7814753 |
| M2.2 | Methampl | 371840 | 324358 | 453980 | 676736 | 1235937 | 398178 | 245340 | 175535 | 144899 | 132470 | 111225 | 111055 | 120249 | 87699 | 100693 | 4690194 |
| M2.3 | Methampl | 396251 | 370201 | 687438 | 1478252 | 3070070 | 801223 | 391653 | 218682 | 174104 | 178991 | 137583 | 128492 | 141110 | 106552 | 149353 | 8429955 |
| M2.4 | Methampl | 419344 | 388211 | 682773 | 1315539 | 2470571 | 737898 | 368597 | 238143 | 186925 | 191016 | 152603 | 143703 | 154617 | 112191 | 157183 | 7719314 |
| M2.5 | Methampl | 403741 | 352812 | 538734 | 998770 | 1786982 | 553071 | 323065 | 200134 | 161223 | 158994 | 129820 | 125689 | 136639 | 91678 | 102950 | 6064302 |
| M2.6 | Methampl | 402997 | 362319 | 665603 | 1468628 | 3481151 | 882655 | 404455 | 204837 | 158864 | 219033 | 142352 | 116353 | 124413 | 91229 | 114711 | 8839600 |
| M2.7 | Methampl | 488019 | 403235 | 567682 | 1178688 | 3188737 | 971553 | 510131 | 273571 | 230784 | 342277 | 218475 | 179694 | 186018 | 142869 | 144708 | 9026441 |
| M2.8 | Methampl | 502697 | 461627 | 809210 | 1396159 | 3054716 | 745410 | 381302 | 257550 | 207327 | 208204 | 173676 | 167310 | 169658 | 136414 | 131880 | 8803140 |
| M2.9 | Methampl | 397053 | 373315 | 671318 | 1996989 | 3658746 | 1277557 | 504024 | 230794 | 180535 | 252391 | 159930 | 139913 | 153577 | 121881 | 146886 | 10264909 |
| M2.10 | Methampl | 437183 | 409539 | 873020 | 1715483 | 2678005 | 746998 | 388261 | 244080 | 195270 | 201955 | 162037 | 147278 | 153618 | 124839 | 142757 | 8620323 |
| M3.1 | Methampl | 819002 | 724017 | 936730 | 1600098 | 2745948 | 976061 | 622869 | 454596 | 392031 | 399414 | 339215 | 336325 | 363685 | 258302 | 267308 | 11235601 |
| M3.2 | Methampl | 899067 | 808225 | 1099107 | 1707278 | 3679889 | 1059948 | 673551 | 457628 | 383536 | 408011 | 321124 | 282630 | 263987 | 211628 | 211876 | 12467485 |
| M3.3 | Methampl | 520234 | 479249 | 764713 | 1556252 | 3391258 | 950862 | 467393 | 265103 | 213469 | 218843 | 174139 | 161717 | 169131 | 130945 | 152900 | 9616208 |
| M3.4 | Methampl | 739626 | 657396 | 748156 | 1046343 | 1997062 | 689590 | 475100 | 357541 | 293719 | 275545 | 231923 | 217382 | 231128 | 171123 | 171301 | 8302935 |
| M3.5 | Methampl | 1178096 | 1046987 | 1298260 | 1849963 | 3438435 | 1127940 | 795184 | 570610 | 480844 | 456012 | 388134 | 350394 | 368298 | 287777 | 284656 | 13921590 |
| M3.6 | Methampl | 480752 | 445352 | 705137 | 1187828 | 2562087 | 638426 | 360403 | 230150 | 198517 | 207551 | 161358 | 137666 | 126307 | 108028 | 112068 | 7661630 |
| M3.7 | Methampl | 730293 | 656614 | 855558 | 1315318 | 2723174 | 808860 | 530168 | 364939 | 307621 | 304759 | 248972 | 229167 | 238923 | 183235 | 179284 | 9676885 |
| M3.8 | Methampl | 649886 | 609564 | 1336311 | 2369219 | 5775278 | 1127320 | 532322 | 306823 | 245338 | 244928 | 191535 | 176607 | 172950 | 144681 | 167260 | 14050022 |
| M3.9 | Methampl | 627001 | 571755 | 778130 | 1271820 | 2606765 | 751453 | 455472 | 324652 | 278785 | 265822 | 225165 | 211541 | 204988 | 169303 | 174732 | 8917384 |
| M3.10 | Methampl | 884689 | 800068 | 1170753 | 2048528 | 3580823 | 1004592 | 601899 | 408742 | 343722 | 327268 | 274041 | 243248 | 257481 | 215650 | 235873 | 12397377 |

**Supplementary Table 8. miRNA with Significant changes in HDPs relative to HCs**

| Gene_ID | Foldchange | p.value | FDR |
|---|---|---|---|
| hsa-miR-375-3p | -1.82 | 2.76E-05 | 1.29E-03 |
| hsa-miR-484 | -1.71 | 8.08E-06 | 6.36E-04 |
| hsa-miR-205-5p | -1.62 | 3.29E-04 | 7.56E-03 |
| hsa-miR-451a | -1.52 | 2.51E-06 | 3.53E-04 |
| hsa-miR-548b-5p | -1.52 | 2.56E-06 | 3.53E-04 |
| hsa-miR-363-3p | -1.47 | 6.15E-08 | 3.39E-05 |
| hsa-miR-132-3p | -1.43 | 2.12E-04 | 5.55E-03 |
| hsa-miR-18a-3p | -1.42 | 1.16E-03 | 0.020 |
| hsa-miR-92b-3p | -1.32 | 7.59E-05 | 2.61E-03 |
| hsa-miR-548c-5p | -1.30 | 1.67E-05 | 1.15E-03 |
| hsa-miR-576-5p | -1.23 | 1.78E-06 | 3.53E-04 |
| hsa-miR-548d-5p | -1.18 | 7.86E-06 | 6.36E-04 |
| hsa-miR-214-3p | -1.14 | 1.65E-04 | 4.98E-03 |
| hsa-miR-1180-3p | -1.12 | 4.54E-05 | 1.79E-03 |
| hsa-miR-92a-3p | -1.09 | 4.68E-06 | 5.16E-04 |
| hsa-miR-3158-3p | -1.08 | 1.92E-03 | 0.028 |
| hsa-miR-181a-5p | -1.04 | 4.37E-05 | 1.79E-03 |
| hsa-miR-7-1-3p | -1.03 | 2.66E-04 | 6.67E-03 |
| hsa-miR-32-5p | -1.01 | 2.43E-05 | 1.29E-03 |
| hsa-miR-486-5p | -0.98 | 2.07E-04 | 5.55E-03 |
| hsa-miR-22-3p | -0.92 | 4.25E-04 | 8.68E-03 |
| hsa-miR-4742-3p | -0.90 | 2.91E-03 | 0.035 |
| hsa-miR-671-5p | -0.88 | 3.86E-03 | 0.043 |
| hsa-miR-101-3p | -0.82 | 1.58E-03 | 0.025 |
| hsa-miR-181d-5p | -0.82 | 2.75E-03 | 0.035 |
| hsa-miR-181b-5p | -0.81 | 2.94E-03 | 0.035 |
| hsa-miR-3150b-3p | -0.78 | 4.43E-03 | 0.048 |
| hsa-miR-16-5p | -0.78 | 1.50E-04 | 4.87E-03 |
| hsa-miR-629-5p | -0.75 | 1.62E-03 | 0.025 |
| hsa-miR-20a-5p | -0.71 | 2.81E-04 | 6.74E-03 |
| hsa-miR-425-5p | -0.70 | 2.82E-05 | 1.29E-03 |
| hsa-miR-4732-3p | -0.64 | 2.70E-03 | 0.035 |
| hsa-miR-15a-5p | -0.63 | 3.60E-03 | 0.041 |
| hsa-miR-21-5p | -0.61 | 1.12E-03 | 0.020 |
| hsa-miR-7-5p | -0.61 | 2.37E-05 | 1.29E-03 |
| hsa-miR-4286 | 0.59 | 2.76E-03 | 0.035 |
| hsa-miR-6514-5p | 0.63 | 1.72E-04 | 4.98E-03 |
| hsa-miR-151b | 0.78 | 9.03E-04 | 0.017 |
| hsa-miR-151a-5p | 0.81 | 3.62E-04 | 7.98E-03 |
| hsa-miR-744-5p | 1.01 | 3.85E-04 | 8.16E-03 |
| hsa-miR-6721-5p | 1.12 | 3.02E-03 | 0.035 |
| hsa-miR-432-5p | 1.23 | 1.57E-03 | 0.025 |
| hsa-miR-191-3p | 1.24 | 1.92E-03 | 0.028 |

**Supplementary Table 9. miRNA with Significant changes in MDPs relative to HCs**

| Gene_id | Foldchange(log2) | p.value | FDR |
|---|---|---|---|
| hsa-miR-375-3p | -1.98 | 3.89E-05 | 3.20E-03 |
| hsa-miR-484 | -1.58 | 4.06E-05 | 3.20E-03 |
| hsa-miR-129-5p | -1.37 | 2.24E-03 | 0.040 |
| hsa-miR-548b-5p | -1.34 | 6.57E-06 | 1.81E-03 |
| hsa-miR-451a | -1.26 | 1.97E-05 | 2.81E-03 |
| hsa-miR-548c-5p | -1.11 | 1.16E-04 | 6.15E-03 |
| hsa-miR-92b-3p | -1.08 | 5.17E-04 | 0.017 |
| hsa-miR-548d-5p | -1.05 | 5.41E-07 | 2.98E-04 |
| hsa-miR-486-5p | -1.02 | 1.34E-04 | 6.15E-03 |
| hsa-miR-363-3p | -0.99 | 2.04E-05 | 2.81E-03 |
| hsa-miR-636 | -0.99 | 1.54E-03 | 0.035 |
| hsa-miR-1180-3p | -0.98 | 1.81E-04 | 7.69E-03 |
| hsa-miR-92a-3p | -0.92 | 4.06E-05 | 3.20E-03 |
| hsa-miR-3150b-3p | -0.87 | 1.82E-03 | 0.035 |
| hsa-miR-4732-3p | -0.86 | 1.34E-04 | 6.15E-03 |
| hsa-miR-5010-3p | -0.85 | 1.78E-03 | 0.035 |
| hsa-miR-629-5p | -0.76 | 1.04E-03 | 0.027 |
| hsa-miR-16-5p | -0.67 | 7.29E-04 | 0.020 |
| hsa-miR-576-5p | -0.66 | 1.86E-03 | 0.035 |
| hsa-miR-4286 | 0.69 | 5.28E-04 | 0.017 |
| hsa-miR-151a-3p | 0.73 | 5.06E-04 | 0.017 |
| hsa-miR-151b | 0.73 | 1.49E-03 | 0.035 |
| hsa-miR-151a-5p | 0.75 | 6.71E-04 | 0.019 |
| hsa-miR-548a-3p | 0.79 | 2.45E-03 | 0.040 |
| hsa-miR-338-3p | 0.81 | 2.50E-03 | 0.040 |
| hsa-miR-744-5p | 0.83 | 1.62E-03 | 0.035 |
